# Characterization of the neoprotein MUC1-fs in patient-derived cells with ADTKD-*MUC1*

**DOI:** 10.64898/2026.09.16.26363125

**Authors:** Karl X. Knaup, Karen Schneider, Hannah Schwarz, René Krüger, Andrea Wenzel, Florian Erger, Charlotte Sachse, Ursula Schlötzer-Schrehardt, Maike Büttner-Herold, Sonja Rehrl, Mario Schiffer, Bruno Hüttel, Bodo Beck, Kathrin Skoczynski, Bjoern Buchholz, Francesca Pasutto, Johannes Schödel, Michael S. Wiesener

## Abstract

Autosomal dominant tubulointerstitial kidney disease (ADTKD) is a genetically heterogeneous group of diseases that regularly lead to kidney failure in mid adulthood. In ADTKD-*MUC1* a distinct neoprotein generated by a frameshift mutation, MUC1-fs, is pathogenic and accumulates in distal renal tubular cells. However, the pathomechanism of the disease and the regulation of the MUC1-fs are poorly understood.

Primary tubular cells from the urine of several probands were established and immortalized. The length of the repeat region of both alleles of *MUC1*, as well as the mutated repeat localization were precisely defined by long range sequencing. The pathogenic MUC1-fs is readily detectable in patient-derived cells and has a considerably longer half-life than the wild-type mucin 1. Targeting the secretory pathway with BRD4780, a compound previously reported as putatively therapeutic, leads to downregulation of MUC1-fs in each of our patient-derived cell models. However, wild-type mucin 1 and numerous other transcriptional pathways were also affected by BRD4780, thus being rather unselective.

Tubular cells derived from the urine appear to be a suitable model to analyze the pathogenic MUC1-fs of an individual patient, enabling pharmacological studies of novel therapies.

## INTRODUCTION

Autosomal dominant tubulointerstitial kidney diseases (ADTKD) are rare, monogenic diseases which lead to kidney failure in mid adulthood by progressive tubulointerstitial fibrosis (Devuyst *et al*, 2019; Eckardt *et al*, 2015). For the two most frequent subtypes of ADTKD (-*UMOD* and -*MUC1*) the altered proteins caused by heterozygous mutations in the germline accumulate in the cytoplasm of the native cells of the tubular apparatus (reviewed in (Devuyst *et al*., 2019; Econimo *et al*, 2022)). It is largely accepted that this form of “toxic proteinopathy” is the mechanism of cellular injury, leading to progressive kidney fibrosis and eventually organ failure. The current literature places both ADTKD subforms (*UMOD* and *MUC1*) into the UPR pathway, which then causes apoptosis (reviewed in (Devuyst *et al*., 2019; Econimo *et al*., 2022)).

Whereas more than 100 distinct *UMOD* variants have been reported to be pathogenic (Devuyst *et al*, 2017), there is only one type of mutation known for ADTKD-*MUC1* leading to a specific frameshift protein (MUC1-fs, neoprotein) in all patients and families known to date (Kirby *et al*, 2013). As such, the majority of patients show a single nucleotide duplication in a highly repetitive domain (variable number of tandem repeats, VNTR), which cannot be detected by standard sequencing techniques and usually requires restriction-specific enrichment and detection by mass spectrometry or SNaPshot minisequencing (Ekici *et al*, 2014; Kirby *et al*., 2013).

In a landmark paper Dvela-Levitt et al. have described the cellular fate of MUC1-fs, being trapped in the early secretory pathway and thereby exerting its toxic effect on tubular kidney cells (Dvela-Levitt *et al*, 2019). Furthermore, they identified the small molecule BRD4780 which binds the cargo receptor Trans-Membrane Emp24 protein transport Domain (TMED9, p24α_2_) in the transport vesicles of the secretory pathway, thereby releasing MUC1-fs and enabling its lysosomal degradation. These data were not only demonstrated in cellular models, but also in a humanized knock-in mouse model for ADTKD-*MUC1*, thus implicating a therapeutical potential of the compound (Dvela-Levitt *et al*., 2019). Since then, not a single independent publication has occurred confirming their crucial findings. Furthermore, the human data therein stem solely from an immortalized cell clone (“P-cell”) derived from a tumor nephrectomy of an ADTKD-*MUC1* patient (Dvela-Levitt *et al*., 2019). As the number of VNTR repeats among individuals varies between 21 and 125 (Gendler *et al*, 1990), the altered protein MUC1-fs of different patients will also vary considerably in length. In addition, the exact repeat position of the mutation is private to each family, thus the ratio of wild-type versus frameshifted repeats will vary between families (Kirby *et al*., 2013; Wenzel *et al*, 2018). Keeping these differences in mind, we aimed to compare and characterize the pathogenic neoprotein from several patients of different families.

Given the generally accepted pathogenic role of cellular MUC1-fs accumulation, therapeutic approaches aimed at downregulating the aberrant protein are likely to confer a clinical benefit. Evaluation of targeted therapeutic strategies against ADTKD-*MUC1* will require cellular models, ideally individualized for different patients. The non-invasive collection and culture of tubular cells from a single void of urine (Zhou *et al*, 2012) appears to be well suited to acquire primary cells from affected individuals, expressing the pathogenic neoprotein MUC1-fs (Knaup *et al*, 2018).

In the current study we established primary tubular cell cultures from numerous ADTKD-*MUC1* patients of different families and immortalized a substantial number of them. We characterized the individual MUC1-fs protein and compared it to the wild-type (wt) mucin 1 in healthy proband’s cells. The secretory pathway inhibitor BRD4780, effectively targets the neoproteins of all patients investigated, but it does not appear to be fully specific.

## METHODS

### Patient and ethical approval

All probands gave written informed consent to all scientific procedures. The studies were approved by the ethics committee of the Friedrich-Alexander University Erlangen-Nürnberg (approval number: 251_18 B).

### Cell culture and immortalization

If not stated otherwise, all reagents were purchased from Sigma-Aldrich (Taufkirchen, Germany). HeLa and MCF-7 cells were supplied from the German Collection of Microorganisms and Cell Cultures (DSMZ, Braunschweig, Germany). HKC-8 cells were a friendly gift from Lorraine Racusen (Racusen *et al*, 1997). The following reagents were used for culture: DMEM, 1 g glucose/L, 10 % fetal calf serum, 2 mmol/L L-glutamine, 100 U penicillin and 100 µg streptomycin/ml (Pen/Strep). Medium as well as penicillin/streptomycin were purchased by PAN-Biotech (Aidenbach, Germany) and fetal calf serum by PAA Laboratories (Coelbe, Germany). Human primary tubular cells (huPTC) were generated and cultured as described by Zhou et al. (Zhou *et al*., 2012). Immortalized Tubular Cells (iTC) were generated by lentiviral SV40 Large T transformation of huPTC using purified lentiviral particles from GeneCopeia (GeneCopoeia, Maryland, USA; Cat No. LPP-SV40LT-Lv153-100-C). Therefore, primary huPTC were seeded in a 24-well plate in DMEM/HAM’s F12 (1:1) (Thermo Fisher Scientific, Massachusetts, USA), supplemented with 100 U/ml Pen/Strep, REGM SingleQuot supplements (cat. no. CC-4127, Lonza, Basel, Suisse), 500 ng/ml amphotericin B (Gibco, Grand Island, New York, USA) and 10 % FCS. The next day cells were infected with 6x10^5^ TU (Transduction Units) lentiviral particles and 8 µg/ml Polybrene in REBM Basal Medium (cat. no. CC-3191, Lonza, Basel, Suisse), supplemented with 100 U/ml Pen/Strep, REGM SingleQuot supplements and 5 % FCS. After 24 h cells were passaged from a 24-well format into a 6-well format and incubated for further two to four days. Then, REBM Basal Medium containing REGM SingleQuot supplements, 100 U/ml Pen/Strep and 2 µg/ml Puromycin (Invivogen, Toulouse, France) was added to initiate the clonal selection process. After further seven to ten days colonies started to form. When these colonies reached a sufficient size they were picked with cloning cylinders (SP Bel-Art, New Jersey, USA) and expanded for further usage.

### Pharmacological treatment of cells

BRD4780 (AGN192403 hydrochloride, Tocris Bioscience; Cat No. 1072) was added in a final concentration of 10 µM for 24 hours, unless otherwise indicated. Cells were exposed to DMOG in a concentration of 1 mM for 18 hours, unless otherwise stated. Cycloheximide (CHX) (Thermo Scientific, Dreieich, Germany) was added at a concentration of 20 µM for the times indicated.

### RNA interference

siRNA knockdowns were performed on all cells over 48h using oligofectamine (Thermo Scientific, Dreieich, Germany) according to the manufacturer’s instructions. All siRNAs were synthetized by Qiagen (Hilden, Germany). See Supplementary Table S2 for details and sequences.

### Transient Transfection

Transient transfections were performed with equal amounts of empty vector (pcDNA3) or frameshift human MUC1-fs plasmid (pcDNA3_MUC1-fs) (Knaup *et al*., 2018) using XtremeGene (Merck, Darmstadt, Germany) transfection reagent according to the manufacturer’s instructions. 24 h after transfection, cells were washed with PBS and either fixed with methanol for immunofluorescent staining or extracted for whole cell protein lysates for immunoblotting.

### RNA extraction

For RNA extraction, cells were homogenized into TRK lysis buffer and extracted via peqGOLD Total RNA Kit (VWR life science, Ismaning, Germany), according to manufacturer’s protocol.

### Protein extraction and immunoblotting

Before harvesting, cells were washed twice with phosphate-buffered saline (PBS, pH7.4) and harvested directly into extraction buffer (8 M urea, 10 % glycerol, 1 % sodium dodecyl sulfate (SDS), 10 mM TrisHCl pH6.8, protease inhibitor complete^TM^ (Roche, Mannheim, Germany) with 500 mM dithiothreitol (DTT; Carl Roth, Karlsruhe, Germany). Lysates were then subjected to ultrasound homogenizer Sonopuls (Bandelin, Berlin, Germany) for 5 seconds. Extracts were then stored at -20°C.

Protein concentrations were measured with the DC Protein Assay (BioRad, California, USA) according to the manufactureŕs instructions. Protein separation was performed using SDS PAGE and proteins were transferred to PVDF membrane (Millipore, Bedford, MA, USA). 5 % milk in TBS-T was used as blocking buffer for 1 h at room temperature after the transfer. For protein detection membranes were incubated with primary antibodies over night at 4°C, followed by an HRP-labeled secondary antibody at room temperature for 60 min (details on antibodies can be found in Supplementary Table S1). Between incubations the membranes were washed 3x5 min using TBS-T and after incubation with the secondary antibody 4x10 min and 2 min in PBS. Signals were visualized by the ECL system (GE Healthcare, Munich, Germany).

### Immunofluorescent staining

#### (A) on cells

Immunofluorescence staining was performed to detect MUC1-fs and TMED9 expression in cultured cells. Cells were seeded on sterile glass coverslips placed in 24-well plates and cultured until reaching approximately 70–80 % confluence. After treatment as required by the experimental design, cells were washed once with PBS (pH7.4) and fixed with ice-cold methanol for 20 min at −20°C. Methanol-fixed cells were subsequently washed three times with PBS to remove residual fixative.

To reduce non-specific antibody binding, cells were incubated in blocking solution composed of 5 % BSA in TBS for 30 min at room temperature. Primary antibody incubation was performed using primary antibodies, as indicated (see Supplementary Table S1 for details). Coverslips were incubated with the primary antibody for 1 h at room temperature in a humidified chamber.

Following three washes with TBS-T (containing 0.1 % Tween-20), cells were incubated with a secondary antibody (see Supplementary Table S1 for details) and DAPI (1 µg/mL) for 45 min at room temperature in the dark. After three additional washes in PBS, coverslips were mounted onto microscope slides using Mowiol (Carl Roth, Karlsruhe, Germany) and dried overnight at room temperature in the dark. Fluorescence imaging was performed using a Leica microscope (Leica DM6000 B).

#### (B) on tissue

Paraffin embedded tissue was cut into 1 µm thick sections and deparaffinized using a decreasing alcohol series. Antigen retrieval was performed by cooking the tissue slides in the microwave for 10 min using 0.1 M citric buffer pH6. After cooling down and three washes with PBS, slides were incubated with a protein block containing 1 % BSA in PBS. Primary antibody staining was performed over night at 4°C after three washes with PBS (see Supplementary Table S1 for details). For secondary antibody binding, slides were washed three times with PBS and incubated for 2 h at room temperature in the dark with goat anti rabbit 488 antibody (see Supplementary Table S1 for details) for MUC1-fs and goat anti mouse 594 antibody for TMED9. After washing three times with PBS, slides were stained with DAPI for 90 sec at room temperature. Slides were mounted with Mowiol and stored in the dark over night before imaging.

### Immunogold Electron Microscopy

For postembedding immunogold labeling, cells were fixed in 4 % paraformaldehyde and 0.1 % glutaraldehyde in 0.1 M cacodylate buffer (pH7.4) for 1 h at 4°C. Cells were dehydrated serially to 70 % ethanol at –20°C and embedded in resin (LR White; Electron Microscopy Sciences). Ultrathin sections were successively incubated in Tris-buffered saline (TBS), 0.05 M glycine in TBS, 0.5 % ovalbumin and 0.5 % fish gelatin in TBS, primary antibodies diluted in TBS-ovalbumin overnight at 4°C, and finally in 10 and 20 nm gold-conjugated secondary antibodies (BioCell, Cardiff, Wales, UK) diluted 1:30 in TBS-ovalbumin for 1 h. After rinsing, the sections were stained with uranyl acetate and examined with a transmission electron microscope (LEO906E, Carl Zeiss Microscopy, Oberkochen, Germany). In negative control samples, the primary antibody was replaced by PBS or equimolar concentrations of nonimmune rabbit IgG or an irrelevant primary antibody.

### RNA-Seq and the Gene Enrichment Analysis (GSEA)

We isolated RNA from iTCs (immortalized tubular cells) using four MUC1-fs positive clones (Kxx) derived from huPTC of three ADTKD-*MUC1*-affected individuals: ADTKD-0150 (K01), ADTKD-0144 (K02), ADTKD-0014 (K17) and ADTKD-0014 (K23). As negative controls, RNA from a MUC1-fs negative clone (K14) from affected individual ADTKD-0014 and a MUC1-fs negative clone from a healthy individual UKER-109 (K03) were analyzed. We used duplicate replicates from cells cultured under control conditions or stimulated with either BRD4780 (10 µM for 24 h) or THP (300 nM for 24 h). The RNA quality of each sample was evaluated using a bioanalyzer (for detailed information, see the Supplementary Information). Libraries were prepared using Novogene’s in-house protocol. The samples were sequenced on an Illumina Novaseq 6000 (Illumina, USA) to a 2x150 paired-end format with an average of 20 million reads per sample (Supplemental Information) by Novogene (Munich, Germany). After a quality check using FastQC (v0.11.8)(Andrews, 2010), the reads were aligned to the human reference genome (hg38) applying the STAR alignment software (v2.6.1c) (Dobin *et al*, 2013). The mapped reads were used to generate a count table using the featureCounts software (version 1.6.1) (Liao *et al*, 2014). Raw reads were filtered by removing reads aligned to Mitochondrial DNA, and the counts of the genes were normalized and visualized using R version 4.1.1. Genes with more than ten counts were retained for further analysis. The DESeq2 package (v1.32.0) was used for logarithmic transformation of the data and data exploration (Love *et al*, 2014). The gene counts were normalized using a variance-stabilized transform (VST). The Principal Component Analysis (PCA) of the VST data was performed using the plotPCA function of the DESeq2 package and visualized using ggplot2 (version 3.3.6) (Wickham, 2011). Differentially expressed genes (DEGs) were identified using the lfcshrink approach in DESeq2. Adjusted p-values were determined using the Benjamini–Hochberg (BH) method within DESeq2. DEGs with an adjusted p-value of ≤0.05 and a log_2_(fold change) ≥ log_2_(0.5) or ≤ –log2(0.5) were defined as upregulated or downregulated, respectively. Gene IDs from the HUGO Gene Nomenclature Committee (HGNC) and Entrez Gene were added to the result files using biomaRT (v2.48.3) (Durinck *et al*, 2009). For each clone of cells we compared RNA-seq data from control conditions to those derived after treatment of the cells with BRD4780 or THP, respectively.

The Gene Set Enrichment Analysis (GSEA) employed 100,000 permutations to define hallmark gene sets enriched in differentially expressed genes from patient-derived cells with MUC1-positive or negative frameshift protein upon treatment with either BRD4780 (10 µM for 24 h) or THP (300 nM for 24 h). This analysis included a weighted enrichment score and a pre-ranking of genes (Subramanian *et al*, 2005). It included all genes with an Entrez gene ID annotation. The analysis used LFC_2_ values (treatment vs control) as the ranking score and the Hallmark gene sets from MsigDB (v7.5.1) to perform enrichment analysis via fgsea (v1.20.0) (https://igordot.github.io/msigdbr/) (Dolgalev, 2020; Korotkevich, 2019). The fgseaMultilevel function was employed, with a minGSSize of 5 and maxGSSize of 800. Adjusted p-values were calculated using the BH approach. Pathways with an adjusted p-value below <0.05 were visualized, and the results files were annotated using org.Hs.eg.db v3.14.0 (Lawrence *et al*, 2013). The results from GSEA were visualized using the ggballoonplot function in the ggpubr package (v0.6.1) (https://rpkgs.datanovia.com/ggpubr/) (Kassambara, 2025).

### UPR activation analysis of RNA-seq data

Raw reads were normalized in R (v4.1.1) using the parameter transcript per million mapped reads (TPM). Gene annotations were added using biomaRt (v2.48.3). The z-score was calculated with base (v4.1.1) via the scale function. Activity of UPR was analysed using predefined marker gene sets of UPR branches (Adamson *et al*, 2016). The z-scores of all genes assigned to one branch were averaged and plotted as the mean value of the respective boxplot using ggpubr (v0.4.0).

## RESULTS

### Characterization of MUC1 VNTRs and immunodetection in proband’s cells

Since the number of repeats in the VNTR of most individuals varies considerably, the respective MUC1-fs protein is private to each family (Kirby *et al*., 2013; Wenzel *et al*., 2018). This is true for the length of the product, as well as the ratio of wild-type (wt) to frameshifted repeats (depending on the position of the mutated repeat). For this reason, we aimed to fully characterize the VNTRs for both alleles by long read single molecule real time (SMRT) sequencing (Vrbacka *et al*, 2026; Wenzel *et al*., 2018; Wenzel *et al*, 2026) for each individual investigated (Table 1).

**Table 1:** Characterization of the proband’s VNTRs and their products of both alleles, respectively.

| **Family** |  |  |  | **A-66** | **A-11** | **A-64** | **A-30** | **A-14** | **A-21** |
| --- | --- | --- | --- | --- | --- | --- | --- | --- | --- |
| **Individual / cell line** | UKER-109 | UKER-108 | MCF-7 | ADTKD-0152 | ADTKD-0014 | ADTKD-0150 | ADTKD-0144 | ADTKD-0061 | ADTKD-0149 |
| **wt VNTR repeats** | 49, 83 | 76, 77 | 44, 73 | 83 | 34 | 46 | n.d. | n.d. | 55 |
| **fs VNTR repeats** | n.a. | n.a. | n.a. | 79 | 69 | 83 | 87 | 60 | 81 |
| **dupC ins position** | n.a. | n.a. | n.a. | 59 | 14 | 28 | 13 | 24 | 18 |
| **wt allele protein size** | 140 kDa, 203 kDa | 196 kDa, 198 kDa | 131 kDa, 185 kDa | 203 kDa | 112 kDa | 126 kDa | n.d. | n.d. | 165 kDa |
| **fs allele protein size** | n.a. | n.a. | n.a. | 158 kDa | 143 kDa | 172 kDa | 180 kDa | 125 kDa | 168 kDa |

Listed in the table are family numbers for ADTKD-*MUC1* affected families (A-XX), with corresponding individual number (ADTKD-0XXX) and healthy probands UKER-108 and UKER-109, as well as the cell line MCF-7. Overall number of wildtype (wt), as well as number of frameshift (fs) VNTR repeats are listed, where determined or applicable. Number of the VNTR repeat, where dupC insertion occurred is listed, where determined or applicable. Aminoacid-specific protein sizes of wt- and fs-allele were predicted by AAT Bioquest (https://www.aatbio.com/tools/calculate-peptide-and-protein-molecular-weight-mw), where applicable. The additional molecular weight change by glycosylation is not considered in the calculations. (n.d., not determined; n.a. not applicable)

We first established human urinary primary tubular cells (huPTC) by a published protocol (Zhou *et al*., 2012) from all probands and performed immunoblotting from huPTC whole cell lysates for the native mucin 1 proteins. We selected the healthy proband UKER-109 who has substantially different lengths of the VNTR (Table 1) and thereby clearly separates both putative allele products by immunoblotting, confirmed with two independent antibodies recognizing distinct epitopes (Figure 1A and 1B). Wherever expression appeared rather weak, we used dimethyloxalylglycin (DMOG), which moderately induces transcriptional regulation of the *MUC1* gene products (Aubert *et al*, 2009; Naas *et al*, 2023), but should not affect the behavior of either protein, mucin 1 or MUC1-fs. The wt-repeats of the VNTR are heavily glycosylated, which greatly exceeds 50 % of the total proteińs molecular mass and can in some instances result in several hundred kilo Daltons, with large tissue dependent differences (Gendler & Spicer, 1995; Hanisch & Muller, 2000; Jensen *et al*, 2010). Thus, the calculated molecular mass for the amino acid sequence of MUC1 (Table 1) does not take the posttranslational modification into consideration, which will substantially retard the migration in an acrylamide gel. Both putative allele products can be markedly reduced by knockdown with MUC1 siRNA (Figure 1C). Another healthy proband, UKER-108, has only one single repeat difference of the two VNTRs of both alleles (76 versus 77 repeats, Table 1). Accordingly, the immunoblot for mucin 1 of her huPTC lysates does not resolve the two allele products and displays a single migrating species (Supplementary Figure S1A).

**Figure 1:**
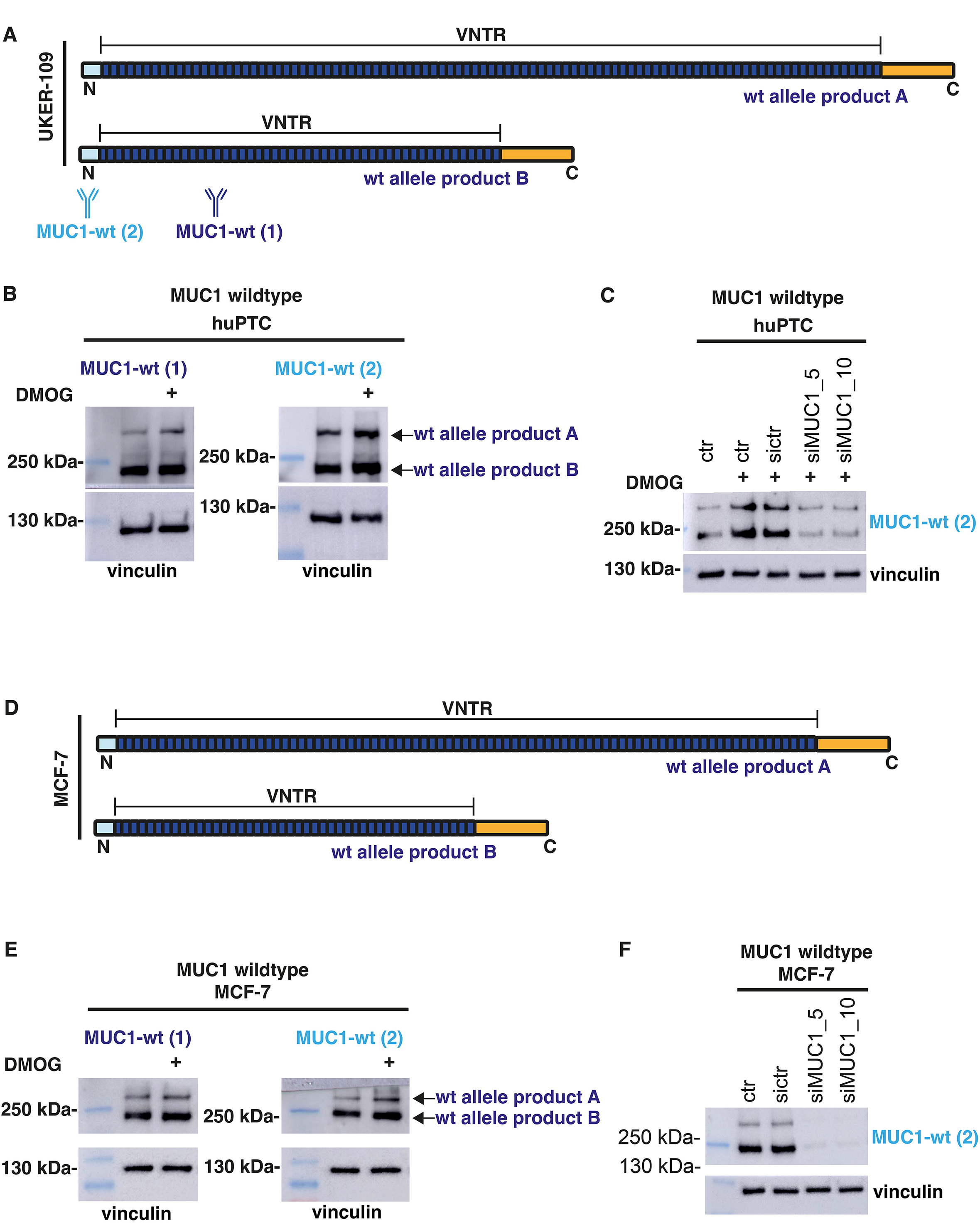
Characterization of mucin 1 in primary cells of a healthy proband and MCF-7 cells. (A) Scheme of both wild-type mucin 1 (MUC1-wt) allele products (A and B) harboring the individual VNTR (Variable Number of Tandem Repeats) domains from proband UKER-109. The exact number of the wildtype repeats within the VNTR are depicted in dark blue. The N-terminal domain (light blue) and the C-terminal domain (orange) is identical in both wildtype mucin 1 allele products. Schematic binding domains of the antibodies used are indicated below. See Table 1 for details on the individual VNTRs. (B) Immunoblots of cell lysates for wt mucin 1 (MUC1-wt (1) and MUC1-wt (2)) from human urinary Primary Tubular Cells (huPTC) from healthy proband UKER-109, corresponding to the scheme in (A). The definite protein sizes are a result of VNTR specific length and mainly glycosylation of the VNTR region. Both antibodies detect the identical *MUC1* allele product. Cells were treated with 1 mM Dimethyloxalylglycin (DMOG) for 18 h to enhance expression levels of mucin 1 through positive transcriptional regulation via the HIF-1-pathway (Naas *et al*., 2023). (C) Immunoblot of cell lysates for MUC1-wt (MUC1-wt (2)) from huPTC from healthy proband UKER-109 exposed to Mucin 1 knockdown by two independent siRNAs (siMUC1_5 and siMUC1_10 or scrambled control (sictr)). Cells were treated with 1 mM DMOG to enhance expression levels of mucin 1 (see B). (D) Scheme of both wildtype allele products harbouring the individual VNTR domains form the mucin 1 expressing cell line MCF.-7 (see (A) for detailed scheme description). (E) Immunoblot of cell lysates from MCF-7 cells showing the two wt allele products (arrows), detected by two independent antibodies: MUC1-wt (1) and MUC1-wt (2). Cells were treated with 1 mM DMOG to enhance expression levels of mucin 1 (see B). (F) Immunoblot of cell lysates for MUC1-wt (MUC1-wt (2)) from MCF-7 exposed to Mucin 1 knockdown by two independent siRNAs (siMUC1_5 and siMUC1_10 or scrambled control (sictr)). See Supplementary Table S1 for details on antibodies. Vinculin serves as loading control in all immunoblots above.

Since the expression of mucin 1 was only moderate even with transcriptional activation with DMOG and the primary huPTC are a timely restricted source for extended repeat experiments, we sought for an immortalized cell line strongly expressing wildtype mucin 1. The carcinoma cell line from the mammary gland MCF-7 fulfilled these criteria. The VNTRs of MCF-7 are also quite different in repeat numbers (44 versus 73 repeats, Figure 1D and Table 1), nresulting in two distinct migrating species upon immunoblotting with both independent antibodies (Figure 1E), which can be profoundly reduced by knockdown experiments (Figure 1F). Having established wildtype mucin 1 immunodetection, we next turned to patient derived huPTC and immunodetection of the neoprotein MUC1-fs. Table 1 shows the results of SMRT sequencing from six ADTKD-*MUC1* patients of different families, with the repeat number of each *MUC1* allele and the exact position of the mutated repeat on the affected allele. Figure 2A shows a scheme of the expected allele products for three families, A-66, -11 and -64. Immunoblots for MUC1-fs of extracts from huPTC of individual patients of these three families display the distinct migration of the MUC1-fs species (Figure 2B), where in most cases two bands were visible, which can again be strongly reduced by RNA silencing (Figure 2C). The anti-MUC1-fs antibody pAb3 was extensively characterized previously, i.e. in patient tissues (Knaup *et al*., 2018), specifically detects MUC1-fs in transfected cells (Supplementary Figure S1B and C) and stains cytoplasmic MUC1-fs in patient’s huPTC but not cells expressing only wildtype mucin 1 (MCF-7, Supplementary Figure S1D), which is predominantly membranous in MCF-7 cells. The quality of the signals is comparable using an antibody directed against the frameshifted VNTR repeats (pAb3-fs, MUC1-fs in Figure 2B, left hand panel), and an antibody specific for the wt-repeats (VU4H5, MUC1-wt (1) in Figure 2B, right hand panel). However, the intensity of the signals corresponds nicely to the respective number of repeats (i.e. product from the A-11 allele with only 34 wt-repeats and 69 frameshifted repeats), implicating multiple binding events of the antibodies. Of note, none of the patient-derived primary cells investigated appeared to express the wild-type protein, which should additionally appear in distinct positions when using the antibodies directed against the wt-VNTR (MUC1-wt (1), Figure 2B) or the N-terminal domain of mucin 1 (MUC1-wt (2), Supplementary Figure 2). This is clearly an *in vitro* phenomenon, as biallelic expression of both alleles is consistently observed in patient kidney tissue (Knaup *et al*., 2018). Interestingly, although the mutated VNTR of family A-64 is larger than that of family A-66, it migrates faster. This is most likely caused by the mutation in a much earlier repeat in the A-64 allele, resulting in many more frameshifted repeats than in the A-66 allele (Figure 2A and Table 1). As described above, the wt-repeats of the VNTR are heavily glycosylated, which profoundly increases the molecular mass of its products. Importantly, this is not the case for the frameshifted repeats (Yamamoto *et al*, 2017).

**Figure 2:**
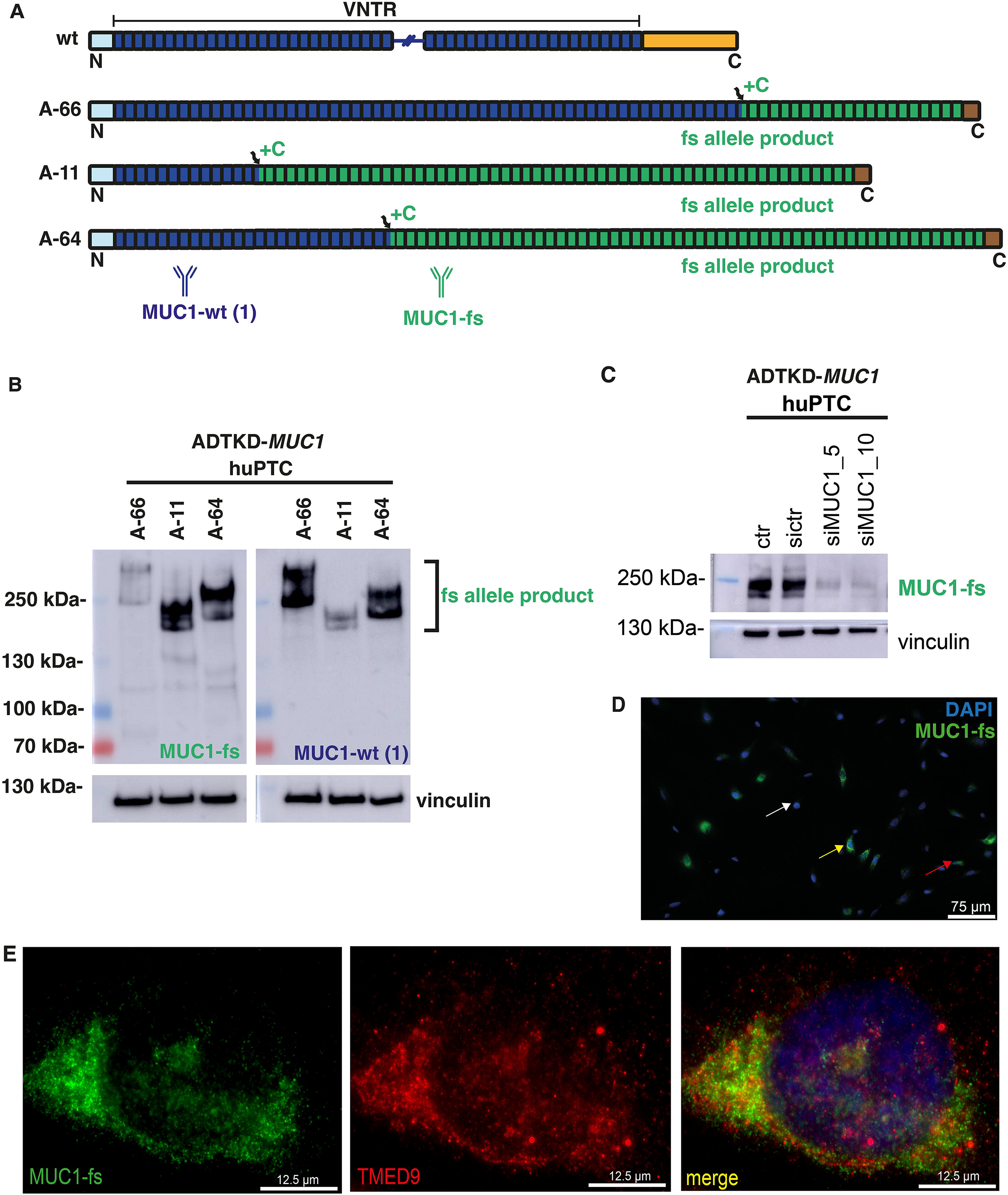
Characterization of MUC1-fs expression in ADTKD-MUC1 patient-derived cells. (A) Scheme of the wild-type mucin 1 (MUC1-wt) protein harboring the VNTR (Variable Number of Tandem Repeats) domain in comparison to the MUC1-fs protein from three ADTKD-*MUC1* affected individuals from families A-66, A-11 and A-64. The exact number of the wildtype repeats within the VNTR are depicted in dark blue, frameshift (fs) repeats in green. The individual positions of the duplication of the additional cytosine base (dupC) within the VNTR, leading to the *de novo* protein MUC1-fs, are indicated by arrows (+C). The N-terminal domain (light blue) is identical in both wildtype mucin 1 and frameshifted MUC1-fs, whereas the C-terminal domain (brown) is *de novo* in MUC1-fs, compared to the wildtype protein (orange in Figure 1A). See also Table 1 for details on the individual VNTRs, position of dupC mutations and predicted protein sizes. Schematic binding domains of the antibodies used are indicated below. (B) Immunoblots of cell lysates for MUC1-fs (MUC1-fs) and wt mucin 1 (MUC1-wt (1)) from human urinary Primary Tubular Cells (huPTC) from the three affected ADTKD-*MUC1* patients, corresponding to the scheme in (A). The definite protein sizes are a result of VNTR specific length and mainly glycosylation of the VNTR region. Both antibodies detect the identical MUC1-fs allele product, as both wt- and fs-repeats are present in the affected allele. Wild-type mucin 1 is obviously not expressed in any huPTC, where additional species would be expected with the MUC1 wt antibody according to the lengths of the wt allele, (see Table 1). Vinculin serves as a loading control. (C) Immunoblot of cell lysates from huPTC from affected individual ADTKD-0149, subjected to MUC1 knockdown by two independent siRNAs (siMUC1_5 and siMUC1_10 or scrambled control (sictr)). Vinculin serves as a loading control. (D) Immunofluorescent staining for MUC1-fs (green) in huPTC from individual ADTKD-0150 from family A-54. Different expression levels of MUC1-fs can be found in the heterogeneous huPTC population, indicated by arrows (yellow: strong, red: weak and white: no expression). Nuclei were stained with DAPI. Magnifications as indicated on scale bars. (E) Immunofluorescent staining for MUC1-fs (green) and TMED9 (red) on huPTC from the affected individual ADTKD-0144 (A-30). The merged image shows a good, but not complete co-localization of both proteins in the cytoplasmic compartment, most likely the early secretory pathway (yellow). Scale bars as indicated.

Immunofluorescent staining for MUC1-fs in huPTCs revealed that only a minority of cells in the primary culture expressed the protein (Figure 2D). This most likely reflects the origin of the tubular cells, as the majority are derived from the proximal tubule (Zhou *et al*., 2012), which does not express mucin 1 (Knaup *et al*., 2018). Among the MUC1-fs–positive cells, expression levels varied markedly (see arrows in Figure 2D). In accordance with previously published data (Dvela-Levitt *et al*., 2019), the cytoplasmic signal for MUC1-fs in the patient-derived huPTC shows accumulation in the cellular secretory pathway, mostly co-localizing with the cargo receptor TMED9 (Figure 2E).

### Effect of inhibition of the secretory pathway with BRD4780

Having established the immunodetection of MUC1-fs from different patients, we next investigated the effect of BRD4780 on several huPTCs. In all cases BRD4780 led to a robust decrease of MUC1-fs (Figure 3A). The effect of BRD4780 can also be appreciated by immunofluorescence, where only a small subset of treated cells exhibited weak cytoplasmic expression of MUC1-fs (Figure 3B).

**Figure 3:**
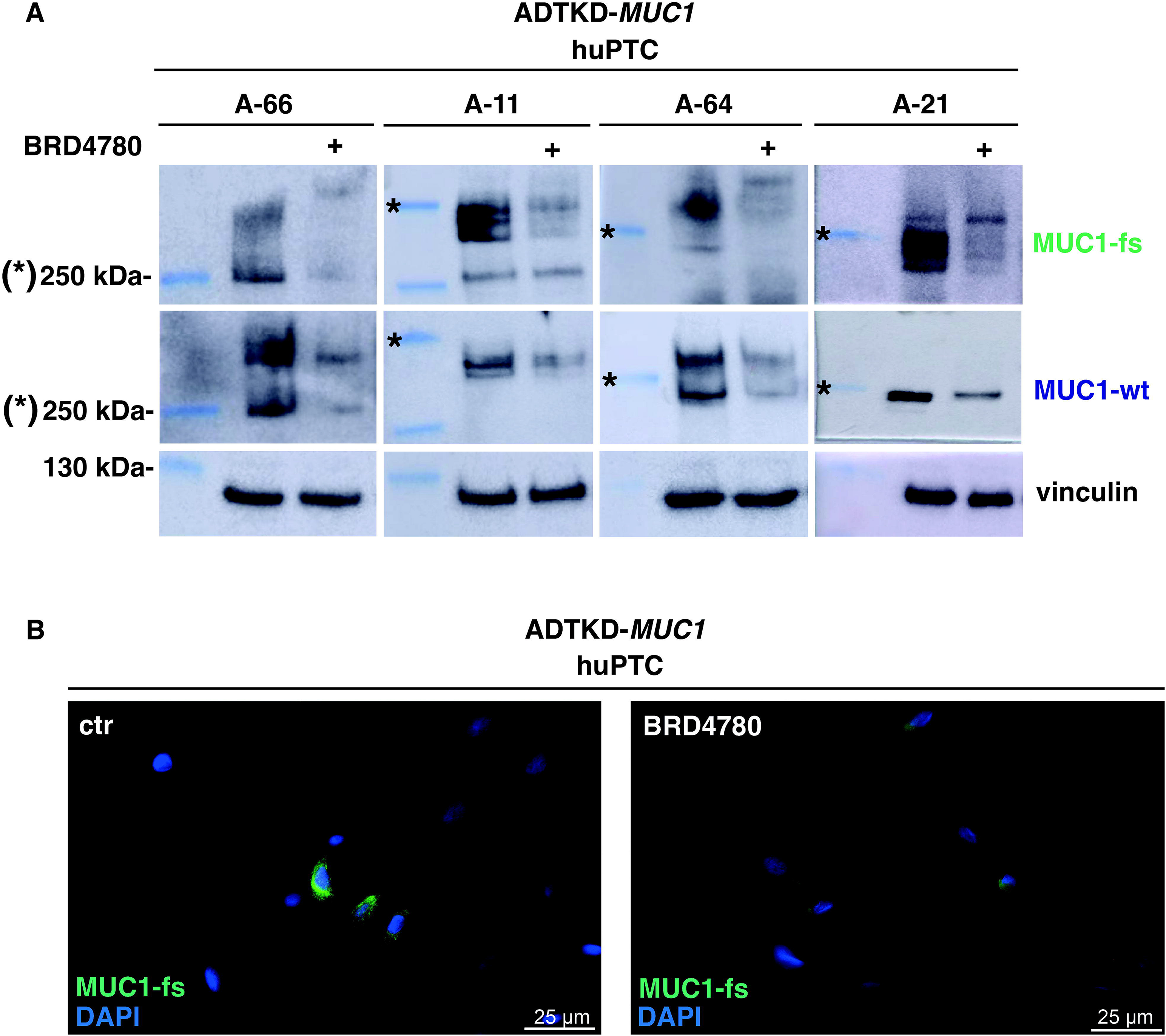
MUC1-fs regulation by BRD4780 in ADTKD-MUC1 patient-derived cells. (A) Immunoblots of cell lysates for MUC1-fs from huPTC of four affected individuals (ADTKD-0152, ADTKD-0014, ADTKD-0150 and ADTKD-0149) from families A-66, A-11, A-64 and A-21, respectively. Cells were treated with BRD4780 (10 µM) for 24 h. MUC1-fs protein is detected with both MUC1-fs and MUC1-wt (1) antibodies, due to the presence of both frameshifted and wildtype repeats within the individual VNTR. Asterisks mark the 250 kDa signal in the respective blots. Vinculin serves as a loading control. (B) Immunofluorescent staining for MUC1-fs (green) in huPTC from the affected individual ADTKD-0150 from family A-64, either control (ctr, solvent) or treated with BRD4780 (10 µM) for 24 h. BRD4780 treatment leads to a clear reduction of cytoplasmic MUC1-fs, yet does not fully eliminate MUC1-fs expression. Nuclear visualization with DAPI. Magnifications as indicated on scale bars.

Since primary cells have experimental limitations, mainly in terms of duration of culture and quantity (i.e. for time course experiments and replicates), we next aimed to immortalize huPTC of numerous patients using viral transformation (Supplementary Figure S3A), generating immortalized Tubular Cells (iTC). Only the cells originating from the distal tubule can be expected to express mucin 1, respectively MUC1-fs. Therefore, we used clonal selection for MUC1-fs expressing cells and favored the clones with stronger expression for further use (Supplementary Figure S3B). BRD4780 also strongly repressed the MUC1-fs in iTC (Figure 4A). As expected, the iTC show a different expression pattern compared to the huPTC in immunofluorescence. Since these are clonal, each cell expresses MUC1-fs, yet again with different expression levels (Figure 4B). Application of BRD4780 leads to a profound downregulation of MUC1-fs in most iTC (Figure 4B, right hand panel). Establishing immortalized clones with expression of MUC1-fs enabled repeat experiments for issues not yet addressed by Dvela-Levitt et al. (Dvela-Levitt *et al*., 2019), such as time course effects of BRD4780 and half-life of the mucin 1 proteins. A time course experiment showed that a strong effect of BRD4780 is already seen after a few hours, with a gradually stronger effect at later time points of continued exposure (Figure 4C), which is paralleled by the effects on TMED9. Cycloheximide time course experiments showed that the half-life of MUC1-fs is about 12 hours (Figure 4D), whereas the half-life of wt mucin 1 protein in MCF-7 cells is only 2-4 hours (Figure 4E), which is similarly short in huPTCs of a healthy proband (Supplementary Figure S4). Thus, the pathogenic neoprotein appears to be at least three times more stable than the wt mucin 1.

**Figure 4:**
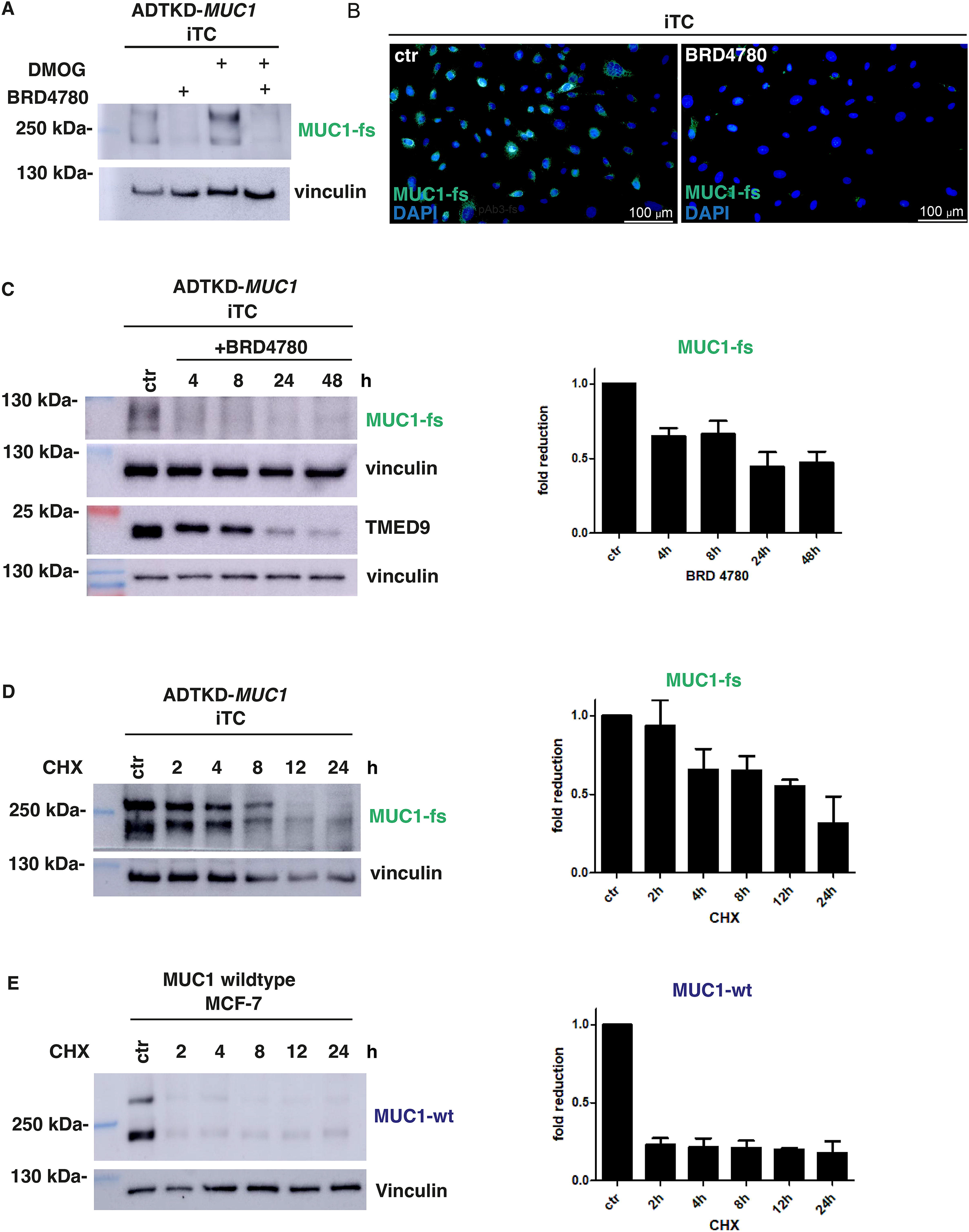
Generation of immortalized tubular cells (iTC) and characterization of MUC1-fs therein. (A) Immunoblot for MUC1-fs of iTC lysates from the affected individual ADTKD-0150 (A-64). iTCs were either treated with solvent, BRD4780 (10 µM) for 24 h, Dimthyloxalylglycin (DMOG, 1 mM to elevate expression levels of MUC1-fs) for 18 h or with both BRD4780 and DMOG. MUC1-fs levels are enhanced with DMOG and reduced by BRD4780 in both settings. Vinculin serves as a loading control. (B) Immunofluorescent staining for MUC1-fs (green) in iTC clone 1 (K1) from the affected individual ADTKD-0150 (A-64), either control (ctr, solvent) or treated with BRD4780 (10 µM, 24 h). BRD4780 treatment leads to a clear overall reduction of cytoplasmic MUC1-fs expression. Nuclei were stained with DAPI. Magnifications as indicated on scale bars. (C) The left panel shows a representative immunoblot of cell lysates for MUC1-fs from iTCs (K2) from the affected individual ADTKD-0144 (A-30). Cells were subjected to BRD4780 (10 µM) for 4, 8, 24 or 48 h. MUC1-fs is clearly reduced already after 4 h of BRD4780 treatment and remains at this low level for up to 48 h. TMED9 shows continuous decrease of expression from 4 h to 48 h. Vinculin serves as a loading control. Quantification of MUC1-fs expression of three independent experiments (n=3) is shown on the right panel with SEM (Standard Error Mean) bars. (D) The left panel shows a representative immunoblot for MUC1-fs of cell lysates from iTCs (K1) from the affected individual ADTKD-0150 (A-64). To determine the half-life of the MUC1-fs protein, iTCs were treated with 20 µM cycloheximide (CHX) for 2, 4, 8, 12 and 24 h. The half-life of the protein is approximately 12 h. Vinculin serves as a loading control. Quantification of MUC1-fs expression of three independent experiments (n=3) is shown on the right panel with SEM bars. (E) The left panel shows a representative immunoblot for mucin 1 of cell lysates from MCF-7 cells. To determine half-life of mucin 1 wildtype protein MCF-7 cells were subjected to CHX (20 µM) for 2, 4, 8, 12 and 24 h. Mucin 1 was detected with MUC1-wt (1) antibody, demonstrating a half-life of the wildtype mucin 1 protein of under 2 h. Vinculin serves as a loading control. Quantification of mucin 1 expression of three independent experiments (n=3) is shown on the right panel with SEM bars.

### Ultrastructural localization of MUC1-fs in iTC

Having established iTCs with expression of MUC1-fs in every single cell, ultrastructural studies on the neoprotein were now possible. Electron microscopy (EM) of iTCs demonstrated often groupwise MUC1-fs immunolabeling in the cytoplasm as well as the endoplasmic reticulum (ER), the former sometimes in vesicle-like structures (Supplementary Figure S5A-B). As expected, the ER did not appear hyperplastic, as reported in murine Umod models (Rampoldi *et al*, 2003) and ADTKD-*UMOD* kidneys (Nasr *et al*, 2008; Reindl *et al*, 2019), an observation which we can also confirm in native kidney biopsies of ADTKD-*MUC1* patients (Supplementary Figure S6). Treatment of cells with BRD4780 greatly reduces immunolabeling of MUC1-fs, with remains of signal being seen in the ER (arrows), whereas knockdown of MUC1 completely clears the ER (Supplementary Figure S5C-D). Double-labeling of iTC with antibodies against MUC1-fs and TMED9, shows that both proteins can be found in the cytoplasm alone (white arrows), as well as together (black arrows), sometimes together in vesicle-like structures (square, Supplementary Figure S5E-F).

### Pleiotropic effects of pharmacological targeting the secretory pathway

TMED9 has been shown to be crucial in cellular MUC1-fs handling (Xiao *et al*, 2024), as well as being the primary target of BRD4780 (Dvela-Levitt *et al*., 2019). We therefore wished to characterize TMED9 and its regulation in our patient-derived cells, as well as kidney tissues. TMED9 is widely expressed in tubules of healthy and ADTKD-*MUC1* kidneys (Figure 5A-C). Co-immunofluorescence demonstrated substantial co-localization of MUC1-fs and TMED9 in human kidney biopsies from ADTKD-*MUC1* patients (Figure 5C). Healthy human kidney displays the apical membranous localization of mucin 1 in distal tubules and no co-localization with TMED9 (Figure 5A), or any signal with an anti-MUC1-fs antibody (Figure 5B). Specific immunoblotting of TMED9 can be verified by knockdown (Supplementary Figure S7A). A strong repression of TMED9 can be achieved with BRD4780 in huPTC and iTC of healthy and affected probands (Figure 5D), as well as the other broadly used cell lines HeLa and HKC-8 (Supplementary Figure S7B). Knockdown experiments for TMED9 in iTCs confirm consecutive downregulation of MUC1-fs (Figure 5E), which is compatible with previously published knockout experiments by CRISPR/Cas9 (Dvela-Levitt *et al*., 2019). Thus, TMED9 appears to be ubiquitously expressed in a wide range of cells and reliably repressed by BRD4780 in all cells. Since TMED9 is a cargo receptor for the general cellular secretory pathway (Strating & Martens, 2009), it cannot be expected to be specific for MUC1-fs. We therefore analyzed other proteins which are known to shuttle and mature through the secretory pathway, randomly selected. Treatment with BRD4780 also led to a decrease of mucin 1 (Figure 6A). On the other hand, the proteins CFTR, ANO1, ANO6, GLUT-1, N-Cadherin and CD71 did not show any difference, at least at the level of whole cell extracts (Supplementary Figure S8). Recently, TMED2 and TMED10 (next to TMED9) have been implicated in binding wildtype and mutant Uromodulin in murine models, as well as being repressed by BRD4780 (Bazua-Valenti *et al*, 2024). We therefore also wished to analyze these proteins for the effect of BRD4780 in our human MUC1-fs model. Unfortunately, the antibody against TMED10 did not appear to be specific judged by immunoblotting and knockdown experiments (data not shown). However, the TMED2 antibody specifically detects its target (Supplementary Figure S9), demonstrating a profound downregulation of TMED2 in iTC and HeLa cells by BRD4780 (Supplementary Figure S9B). Interestingly, the exposure to BRD4780 shows a very similar time-course effect on TMED2 and TMED9 (compare Supplementary Figure S9C to Figure 4C). We next investigated the effect of BRD4780 on the transcriptome and compared this to the effect of Thapsigargin (THP), which is a strong inducer of the unfolded protein response (UPR; Figure 6B) in different iTC. Rather unexpected, in a Gene Set Enrichment Analysis (GSEA) using differentially regulated genes and the Hallmark gene sets (MsigDB) BRD4780 on its own led to robust induction of the UPR signal across all cell-lines, which was similar to that induced by THP. Importantly, this effect was independent from the presence of MUC1-fs expression. In addition, BRD4780 (but not THP) induced targets involved in protein secretion, which may reflect a counter-regulation to the repression of TMEDs. BRD4780 caused multiple further effects in different pathways, which seemed to be much more cell- and context dependent than exposure to THP (Figure 6B and Supplementary Figure S10). Taken together, BRD4780 leads to pleiotropic cellular responses beyond MUC1-fs downregulation, which may lead to undesired effects in a living organism.

**Figure 5:**
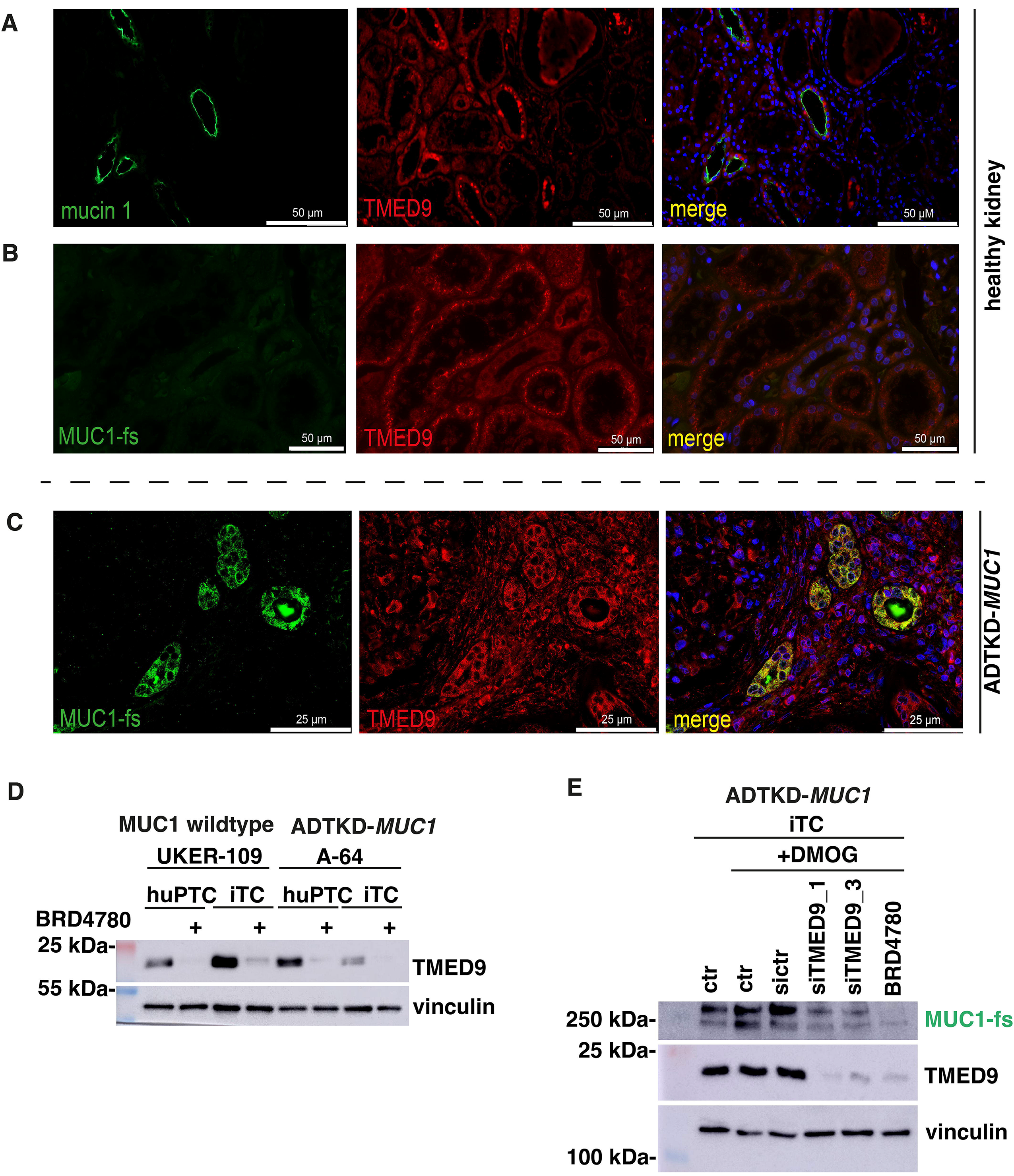
Expression and regulation of the cargo receptor TMED9. (A) Immunofluorescent staining of mucin 1 (green) and TMED9 (red) on tissue of a healthy kidney. The merged image shows no co-localization of both proteins. Magnification as indicated by scale bars. (B) Immunofluorescent staining for MUC1-fs(green) and TMED9 (red) in healthy kidney. Only TMED9 shows tubular protein expression, whereas MUC1-fs is completely negative in tissue of a healthy kidney. (C) Immunofluorescent staining for MUC1-fs (green) and TMED9 (red) on kidney tissue of the affected individual ADTKD-0034 (A-29) (Knaup *et al*., 2018). The merged image shows strong co-localization (yellow) of both proteins in MUC1-fs expressing tubules. Magnification as indicated by scale bars. (D) Immunoblot for TMED9 of cell lysates from huPTC and iTCs of the healthy proband UKER-109 and the affected individual ADTKD-0150 (A-64), comparing BRD4780 treated cells (10 µM for 24 h) to controls. TMED 9 expression levels are clearly reduced in all BRD4780 treated cells. Vinculin serves as a loading control. (E) iTCs from affected individual ADTKD-0150 (A-64) were subjected to siRNA knockdown of TMED9, using two commercially available siRNAs (siTMED9_1 and siTMED9_3). TMED9 levels are markedly decreased after knockdown and also after treatment with BRD4780 (10 µM) for 24 h. An induction of MUC1-fs is observed after DMOG treatment and both TMED9 knockdown and BRD4780 treatment effectively reduce MUC1-fs levels. Vinculin serves as loading control.

**Figure 6:**
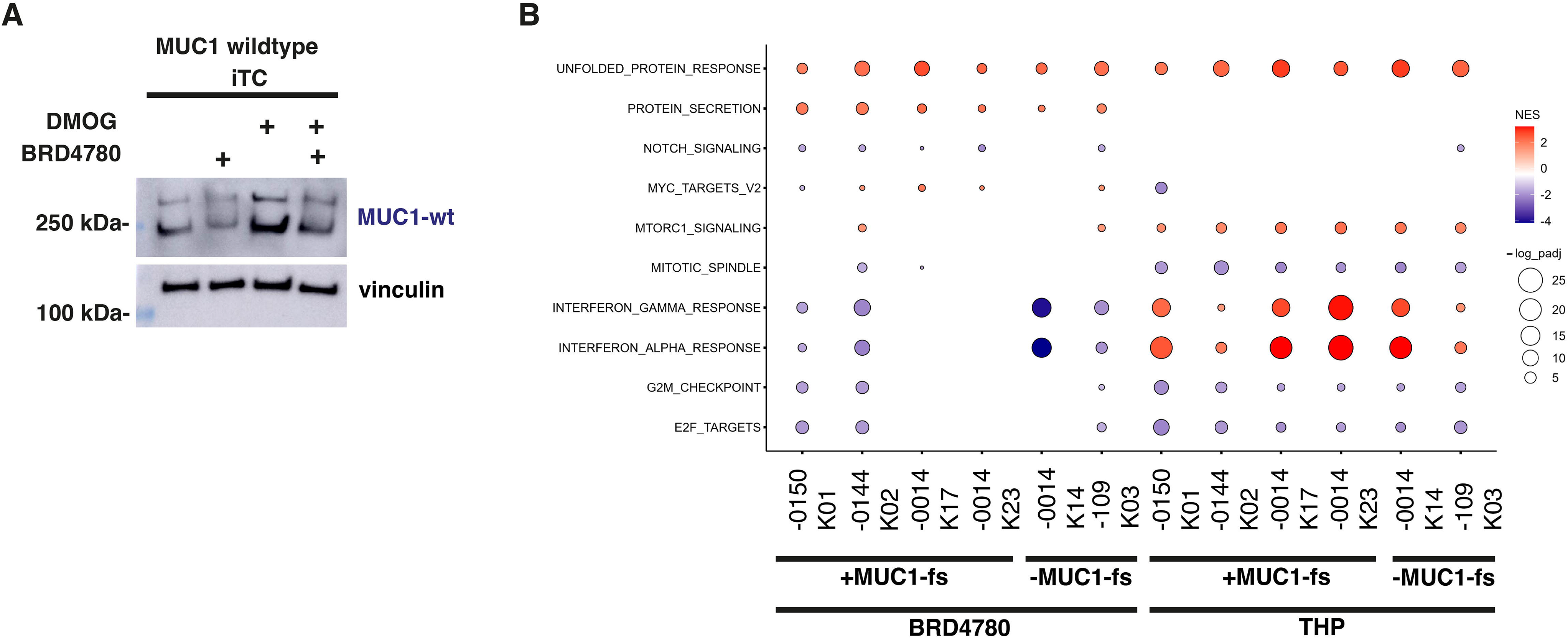
Broad effects of blocking the secretory pathway. (A) Immunoblot of cell lysates from healthy proband UKER-109 for mucin 1. Cells were treated with BRD4780 (10 µM) for 24 h. DMOG (1 mM) was added for 18 h. The two individual MUC1 allele products are detected with the MUC1-wt (1) antibody and correspond to the calculated protein sizes (see Figure 1 and Table 1). BRD4780 leads to a decrease of mucin 1 protein levels, whereas DMOG increases basal levels of the protein. Vinculin serves as a loading control. (B) Bubble plot showing significantly enriched hallmark gene sets from MsigDB determined by Gene Set Enrichment Analysis (GSEA) using RNA-seq data from the indicated clones of cells. Comprehensive RNA sequencing was conducted on iTCs derived from five distinct clones (denoted as Kxx) originating from three affected individuals diagnosed with ADTKD-*MUC1*, specifically ADTKD-0150 clone 1 (K01), ADTKD-0144 (K02), and ADTKD-0014 (K17 and K23). As negative controls, we incorporated a MUC1-fs negative clone (K14) sourced from ADTKD-0014 and a clone (K03) from a healthy individual (UKER-109; also MUC1-fs negative). These samples were subjected to RNA-seq analysis utilizing duplicate replicates. Cells were cultured under controlled conditions with either 10 µM BRD4780 for 24 h or 300 nM THP for 24 h, respectively. Differential expression analysis was conducted for each cell clone comparing either BRD4780 or THP treatment to control conditions. Enrichment of MsigDB hallmark gene sets was determined by GSEA based on an adjusted p-value threshold of <0.05, employing the Benjamini-Hochberg correction method. Gene sets were subsequently visualized according to normalized enrichment scores (NES) and - log10 (adjusted p-values). Red color indicates pathways that were induced by the respective treatment, while blue color depicts downregulated pathways. Only the pathways identified consistently across all four MUC1-fs positive clones per treatment are presented in this figure. A detailed overview of all significant hallmark gene sets identified in this study is available in Supplementary Figure S10.

## DISCUSSION

Rare kidney diseases account for an extremely heterogeneous group of conditions and pose an increased risk for kidney failure as compared to all cause chronic kidney disease (CKD) (Wong *et al*, 2024). The majority of rare kidney diseases are thought to be hereditary (Devuyst *et al*, 2014), with more than 600 genes being implicated in Mendelian kidney diseases (Rasouly *et al*, 2019). Nevertheless, the number of truly actionable genes in clinical nephrology is painfully restricted to date (KDIGO, 2022). This is most likely due to the lack of pathomechanistic understanding of most Mendelian kidney diseases, which impairs the development of effective, targeted therapies.

The first patients with ADTKD may have been reported in 1962, then termed “medullary cystic disease” (Strauss, 1962). The corresponding disease for ADTKD-*MUC1* was first linked to a locus on chromosome 1q21 in 1998 (Christodoulou *et al*, 1998). However, because of the complexity of *MUC1*, identification of the causative gene was delayed until 2013 (Kirby *et al*., 2013). The unique neoprotein MUC1-fs caused by characteristic mutations in or upstream of the VNTR of *MUC1* is clinically readily detectable in kidney samples of ADTKD-*MUC1* patients and thought to be disease-causing (Knaup *et al*., 2018; Zivna *et al*, 2018). The observed cytoplasmic accumulation of MUC1-fs has been largely resolved to be mainly in compartments of the early secretory pathway, in particular in Coat protein (COP) I and II vesicles, with an affinity to the cargo receptor TMED9 (Dvela-Levitt *et al*., 2019). Importantly, this study has also identified a small molecule, BRD4780, which regulates TMED9 and thereby releases MUC1-fs from the early secretory pathway towards lysosomal degradation, thus being positioned as a potential therapeutic agent (Dvela-Levitt *et al*., 2019).

Our study is the first to confirm the effect of BRD4780 in downregulating MUC1-fs in a range of patient-derived cells. The onset of this effect is faster than the half-life of the protein (compare Figure 4C to D), which is compatible with active degradation of MUC1-fs as a consequence of the treatment with BRD4780. Interestingly, the half-life of wild-type mucin 1 is shorter than that of mutated MUC1-fs (compare Figure 4E to D, respectively), which may reflect the cytoplasmic accumulation of the neoprotein. However, the short-term effects on a cellular level may not be as obvious and possibly not relevant for an *in vivo* situation on a tissue level, since matured mucin 1 integrated into the membrane will not be targeted by BRD4780 (the same being possible for other proteins investigated, as shown in Supplementary Figure S8).

The secretory pathway is a broad route for maturation of about a third of all synthetized proteins of any cell (Gomez-Navarro & Miller, 2016; Malhotra, 2025). The cargo receptors of the TMED group are strongly involved in the shuttling machinery (Strating & Martens, 2009), and therefore not specific for any given protein (such as mucin 1 or MUC1-fs). As shown previously (Strating & Martens, 2009), we can confirm broad expression of TMED9 in kidney tissue and a range of primary and immortalized cells (Figure 5 and Supplementary Figure S7), as well as strong repression by BRD4780 (Figure 5D and Supplementary Figure 7B) in all these backgrounds. Interestingly, another TMED, TMED2, is also profoundly repressed by BRD4780 (Supplementary Figure S9), which further increases our notion that intervening at this point may cause off-target effects and possibly side effects for an organism. The RNAseq performed in our patient-derived cells probably indicates such effects by BRD4780 (Figure 6B and Supplementary Figure S10), where a number of pathways show significant regulation by BRD4780, yet seemingly independent of the cellular status of MUC1-fs. Interestingly, BRD4780 itself induced the UPR pathway in our cells, which was comparable to the established UPR inducer THP. Rather unexpected, BRD4780 also induced the pathway for protein secretion, which may be a counter-regulation resulting from pharmacological inhibition of the secretory pathway. Importantly, in our experiments UPR was also not altered by the MUC1-fs status of the cells (Supplementary Figure S10), as well as forced expression of MUC1-fs (data not shown). Therefore, in our view more work needs to be invested in clarifying the molecular mechanism(s) of ADTKD-*MUC1*.

Another effective and specific method to silence a specific gene could be targeting by RNA interference. Until today, an increasing number of oligonucleotides are approved for therapy of different diseases, so far with good safety data (Ebenezer *et al*, 2025; Ghasemiyeh & Mohammadi-Samani, 2025). A disease caused by a dominant negative mutation, such as ADTKD-*MUC1* (or -*UMOD*) is probably an ideal objective for oligonucleotide targeting since silencing of the altered gene product should be therapeutic. As expected, knockdown with siRNA directed against different sequences of MUC1 leads to a convincing reduction of the pathogenic MUC1-fs (Figure 2C). However, the wild-type protein would also be affected, since the siRNA cannot differentiate between the two alleles (Figure 1 C and E). Knockout studies in mice have shown that loss of mucin 1 is not deleterious in (unstressed) mice (Spicer *et al*, 1995). However, whether this is also true in humans is not known to date.

To date, the primary target organ of approved oligonucleotides is the liver (Ebenezer *et al*., 2025; Ghasemiyeh & Mohammadi-Samani, 2025). The kidney as primary target for RNA interference has so far not been developed, yet many authors recently discuss its utility (Ahn *et al*, 2023; Bondue *et al*, 2023; Hu *et al*, 2025). Circulating naked siRNA are freely filtered by the glomeruli and show a very high kidney tissue concentration (Geary *et al*, 2015; Gokirmak *et al*, 2021). Knockdown effects are strong in the proximal tubular system, but only weak in the distal tubule (Donner *et al*, 2018), where oligonucleotides against ADTKD-*MUC1* would need to be directed against (Knaup *et al*., 2018). Thus, drug delivery may need to be developed towards the distal tubule.

An obvious limitation of our study is the exclusive use of cell culture models. The fact that the cells derived from ADTKD-*MUC1* patients do not seem to express the wild-type protein is clearly a cell culture phenomenon. Therefore, some of our data concerning regulation of wild-type mucin 1 may probably not translate to the situation *in vivo*. However, the focus of our study was to learn more about expression and regulation of the pathogenic neoprotein MUC1-fs, for which we used primary cells of several patients. These data in aggregate and possibly this cellular method should be valuable for development of any therapeutic strategy.

In conclusion, the neoprotein in ADTKD-*MUC1* is a plausible and druggable target for therapeutic intervention. Blocking the secretory pathway may face the challenge of being too broad and cause side-effects. RNA interference may be more specific but will require establishment of efficient delivery to the distal renal tubule. In the interest of development of an effective and targeted treatment in our view both avenues deserve further development.

## DATA AVAILABILITY

Data generated for this project have been deposited in the GEO database (https://www.ncbi.nlm.nih.gov/geo/) under accession number GSE306788. All other data that support the findings of this study are available in the Materials and Methods, Results, and/or Supplemental Material of this article.

## AUTHOR CONTRIBUTIONS

KXK and MSW performed conceptualization, data curation, funding acquisition and wrote the original draft. KS, HS, RK, FE, CS, USS, MB, SR, MS, BH, BB, KS, BjB, FP and JS performed partial investigations. All authors reviewed and approved the final version of the manuscript.

## ACKNOWLEDGEMENTS

We would like to thank Yvonne Thoss for her valuable work on the patient’s registry and Daniela Schweitzer for excellent technical assistance. The authors wish to express their gratitude to all patients who generously contributed to this study and the family alliance “ADTKD *Vision Cure*” (www.adtkd.de). This work was partially performed in fulfillment of the requirements for obtaining the degree Doctor of Medicine (Dr. med.) by CS and SR.

