## Supplementary Data for "Characterization of the neoprotein MUC1-fs in patient-derived cells with ADTKD-*MUC1*"

<u>Table of contents</u>:

1. Supplementary Figures and Figure Legends S1-S10 (pages 2-19)

2. Supplementary Table S1: Antibodies applied in our study (page 20)

3. Supplementary Table S2: siRNAs applied in our study (page 21)

4. Supplementary References (page 22)

Supplementary Figure S1

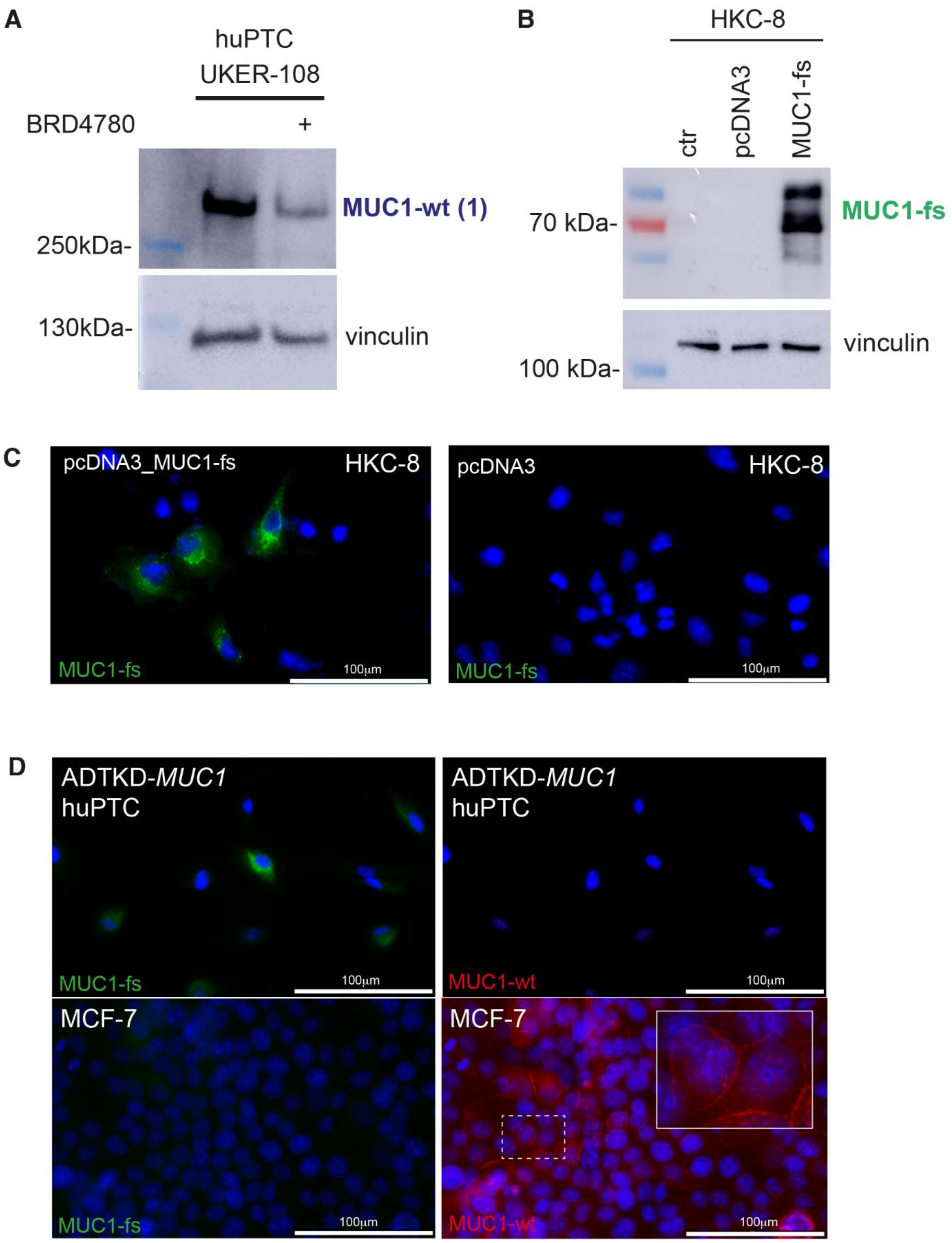

**Supplementary Figure S1:** *Antibody specificity*

(A) Immunoblot of whole cell lysates for Mucin 1 (MUC1-wt (1) antibody) of huPTC from healthy proband UKER-108. Mucin 1 expression level was reduced after treatment with BRD4780 (10 mM) for 24 h. The two VNTRs of this proband are only one repeat apart (76 versus 77 repeats; see Table 1). Therefore, the immunoblot only resolves one single band. Vinculin serves as loading control. (B) Immunoblot of whole cell lysates for MUC1-fs of HKC-8 cells after transient transfection with either pcDNA3 or pcDNA3_MUC1-fs. Bands are only visible in cells transfected with pcDNA3_MUC1-fs. Vinculin serves as loading control. (C) Immunofluorescent staining for MUC1-fs (green) after transfection with either pcDNA3 (no signal) or pcDNA3_MUC1-fs (encoding full length MUC1-fs cDNA; clear cytoplasmic accumulation). DAPI was used for nuclear visualization. Scale bars as indicated. (D) Immunofluorescent staining for MUC1-fs (green) of huPTC from affected individual ADTKD-0150 (A-64) (upper panel) shows clear cytoplasmic signal in cells most probably deriving from the distal tubule. Mucin 1 (red) expression is lost in all huPT cells. Lower panel shows clear positive immunofluorescent staining for mucin 1 in MCF-7 cells (lower right panel), mostly located in the membrane (digitally enlarged section). No MUC1-fs expression can be detected in MCF-7 cells. Scale bars as indicated.

Supplementary Figure S2

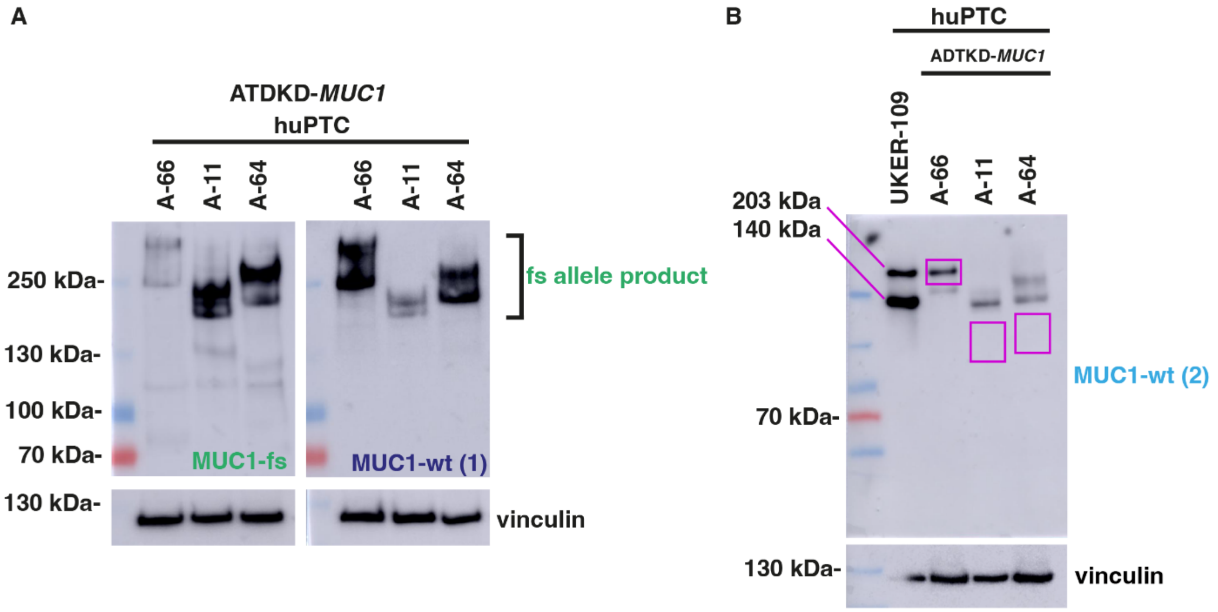

**Supplementary Figure S2:** *Analysis of different Mucin 1 / MUC1-fs epitopes*

(A) Repeat illustration (Figure 2B) of immunoblots from lysates of huPTCs of three ADTKD-*MUC1* patients for MUC1-fs with the antibody pAb3-fs (left hand panel). The antibody against wild type repeats of the MUC1 VNTR (MUC1-wt (1), right hand panel) detects the MUC1-fs, but should also detect the second, non-mutated allele product (if expressed). In most cases this would migrate at a different position (see Table 1). (B) The very same lysates of ADTKD-*MUC1* patients were blotted with an independent antibody against wild-type mucin 1, yet detecting the N-terminal region of the protein (MUC1-wt (2)). The lysates of the huPTCs from the healthy proband UKER-109 display the two mucin 1 wild type allele products defined in Figure 1A and 1B, corresponding to the calculated molecular weights of 203 kDa and 140 kDa, respectively. Of note, as described in the Results section each repeat of the wt-VNTR is heavily glycosylated, which substantially increases its molecular weight and retards its migration in an acrylamide gel. In search of the wild-type allele products of the ADTKD-*MUC1* patients the fully glycosylated mucin 1 allele products of the healthy proband UKER-109 serve as a size control (indicated on the left). The predicted positions of the respective wild-type allele products of the patient derived cell lysates are indicated in pink bars (calculated molecular weight [Table 1] plus glycosylation). No additional signal corresponding to the wild-type allele products can be detected with either antibody.

Supplementary Figure S3

A

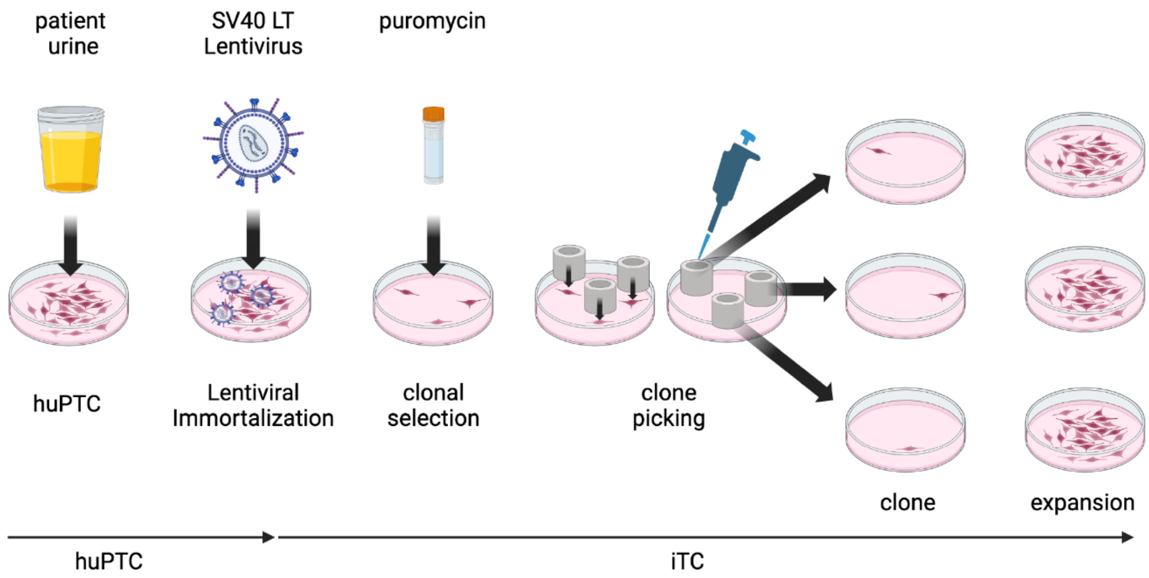

B

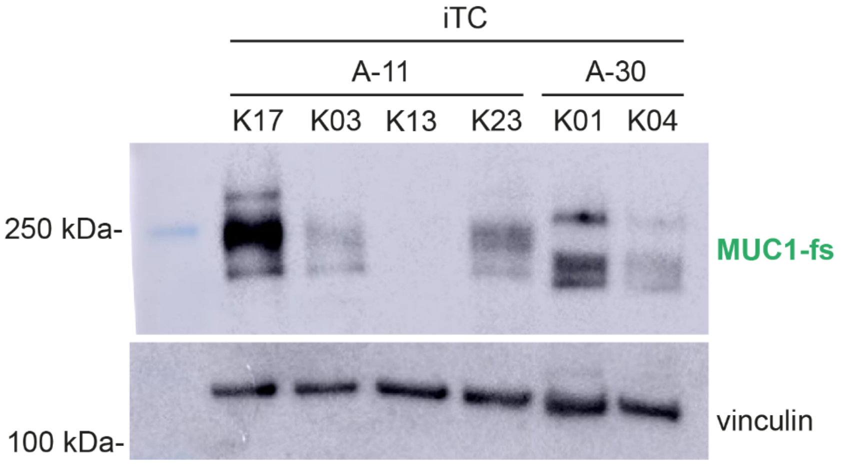

**Supplementary Figure S3:** *Generation of immortalized tubular cells (iTCs)*

(A) Graphical illustration (BioRender) of the established pipeline to generate immortalized Tubular Cells (iTCs) via lentiviral SV40 Large T-Antigen transformation. The primary step is to generate huPTC from the urine of patients or healthy controls, following the protocol from Zhou et al.(Zhou *et al*, 2012). Once huPTC are established, they are infected with SV40 LT Lentivirus according to manufacturer's protocol (GeneCopoeia). After clonal selection with puromycin (2 µg/ml), iTC cellular clones can be picked using cloning cylinders. iTC clones are then expanded for further analysis. (B) Immunoblot analysis of MUC1-fs expression in individual iTC clones (Kxx) generated from huPTC of two families (A-11 and A-30) from affected individuals ADTKD-0014 and ADTKD-0144, respectively. Clones K17, K03 and K23 are positive for MUC1-fs, whereas K13 of the same family is negative for MUC1-fs expression. Positive clones most likely have a distal tubular origin, compared to a non-distal origin of negative clones. Clones K01 and K04 of family A-30 are both positive for MUC1-fs expression. Vinculin serves as loading control.

Supplementary Figure S4

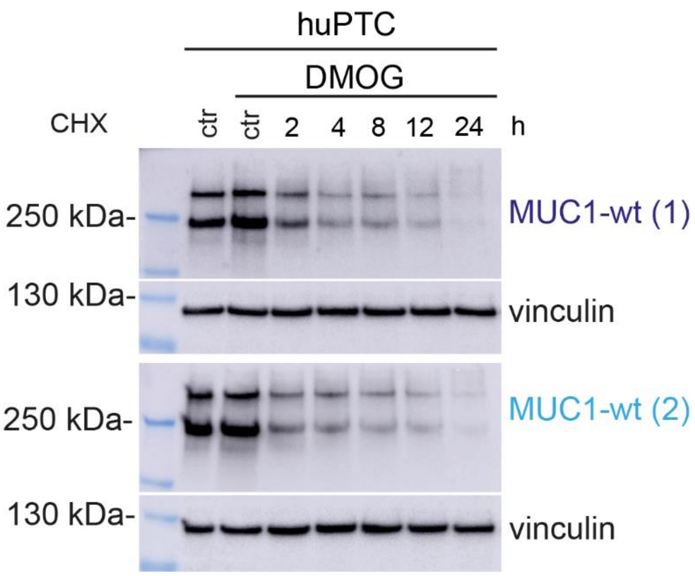

**Supplementary Figure S4:** *Protein half-life evaluation of mucin 1*

Immunoblot for mucin 1 of cell lysates from healthy control huPTC from individual UKER-109. To determine half-life of mucin 1 wildtype protein huPTC were subjected to CHX (20 µM) for 2, 4, 8, 12 and 24 h. DMOG (1 mM) was added to induce the levels of MUC1-fs and two individual mucin 1 antibodies were used for detection (MUC1-wt (1) and MUC1-wt (2)). Both antibodies produce similar results, demonstrating a half-life of the wildtype mucin 1 protein of about 2 h. Vinculin serves as a loading control.

Supplementary Figure S5

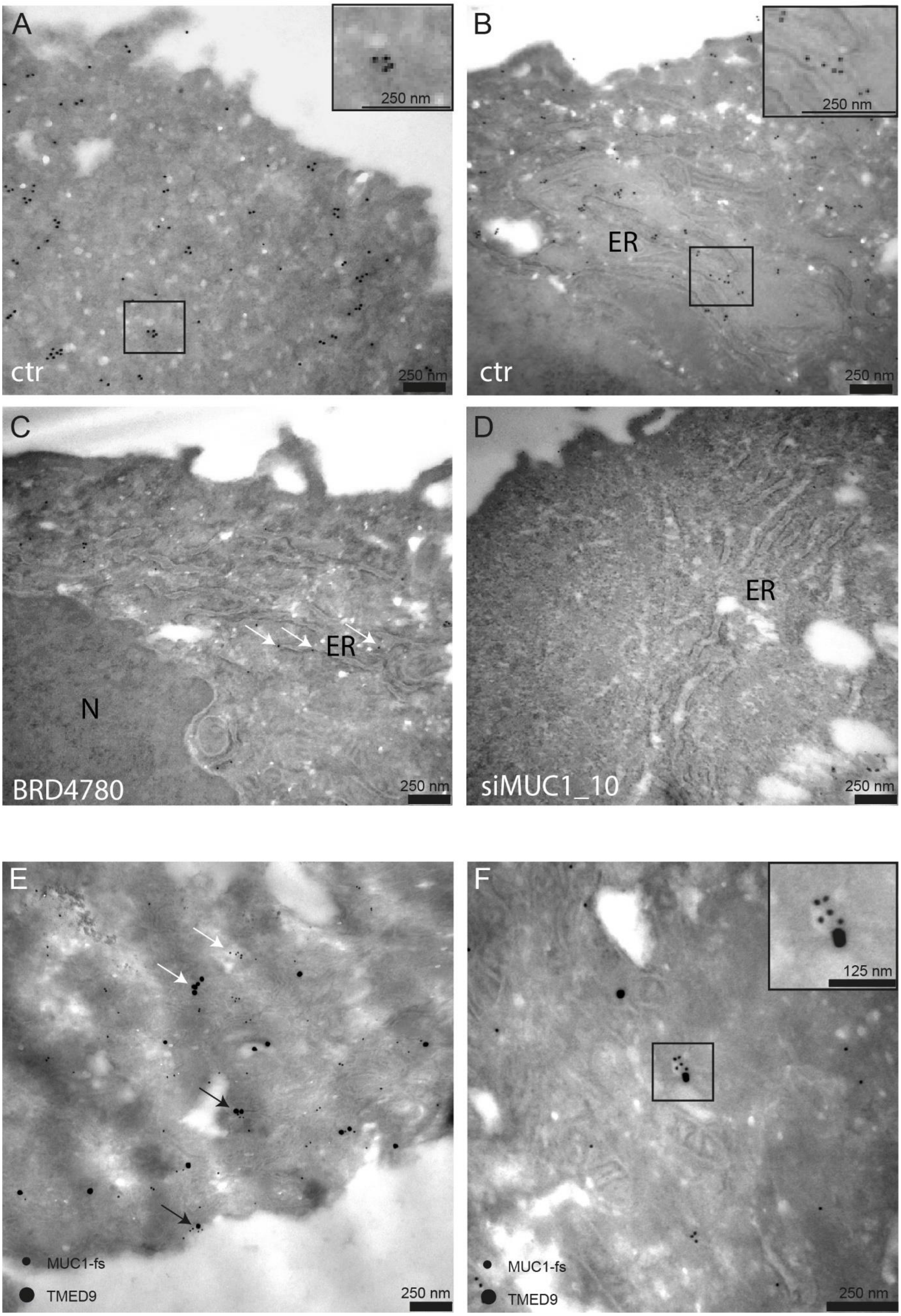

**Supplementary Figure S5:** *Immunogold electron microscopy of MUC1-fs and TMED9*
(A) Immunogold electron microscopy for MUC1-fs in iTCs (K1) from the affected individual ADTKD-0150 (A-64) shows partial aggregation of MUC1-fs protein in vesicular structures and within close vicinity (B) of the endoplasmatic reticulum (ER), as depicted in the digital enlargements of the marked areas in the upper right corners of (A) and (B). (C) After treatment with BRD4780 (10 µM) for 24 h, overall MUC1-fs levels are clearly reduced, yet remain visible within the ER, as indicated by white arrows. The nucleus (N) remains clear of MUC1-fs protein, as expected. (D) After siRNA knockdown of MUC1 (siMUC1_10) an overall clear reduction of MUC1-fs protein can be seen, also within the ER. (E) Immunogold double staining of MUC1-fs and TMED9 reveals isolated signals of MUC1-fs and TMED9 (white arrows), as well as localization of both proteins in close vicinity (black arrows) (MUC1-fs particle size: 10 nm; TMED9 particle size: 20 nm), predominantly in vesicular structures (F) outside of the ER. Magnifications as indicated on scale bars.

Supplementary Figure S6

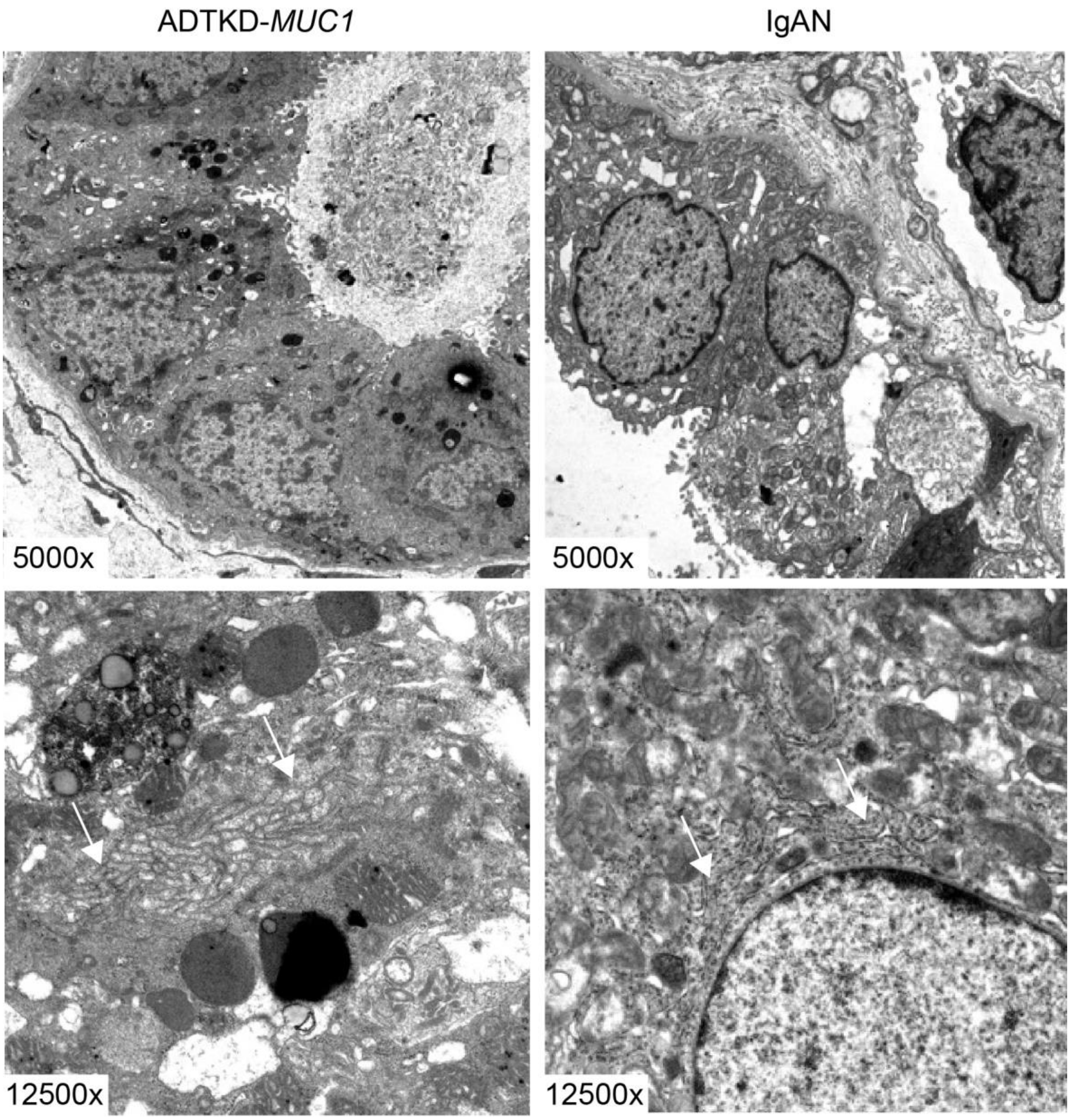

**Supplementary Figure S6:** *Regular appearance of the endoplasmic reticulum in ADTKD-MUC1*

Electron microscopy of kidney sections of an ADTKD-*MUC1* patient (left panel) and an IgAN (IgA Nephropathy) patient (right panel) showing physiological appearance of the Endoplasmic Reticulum (white arrows). Magnifications as indicated.

Supplementary Figure S7

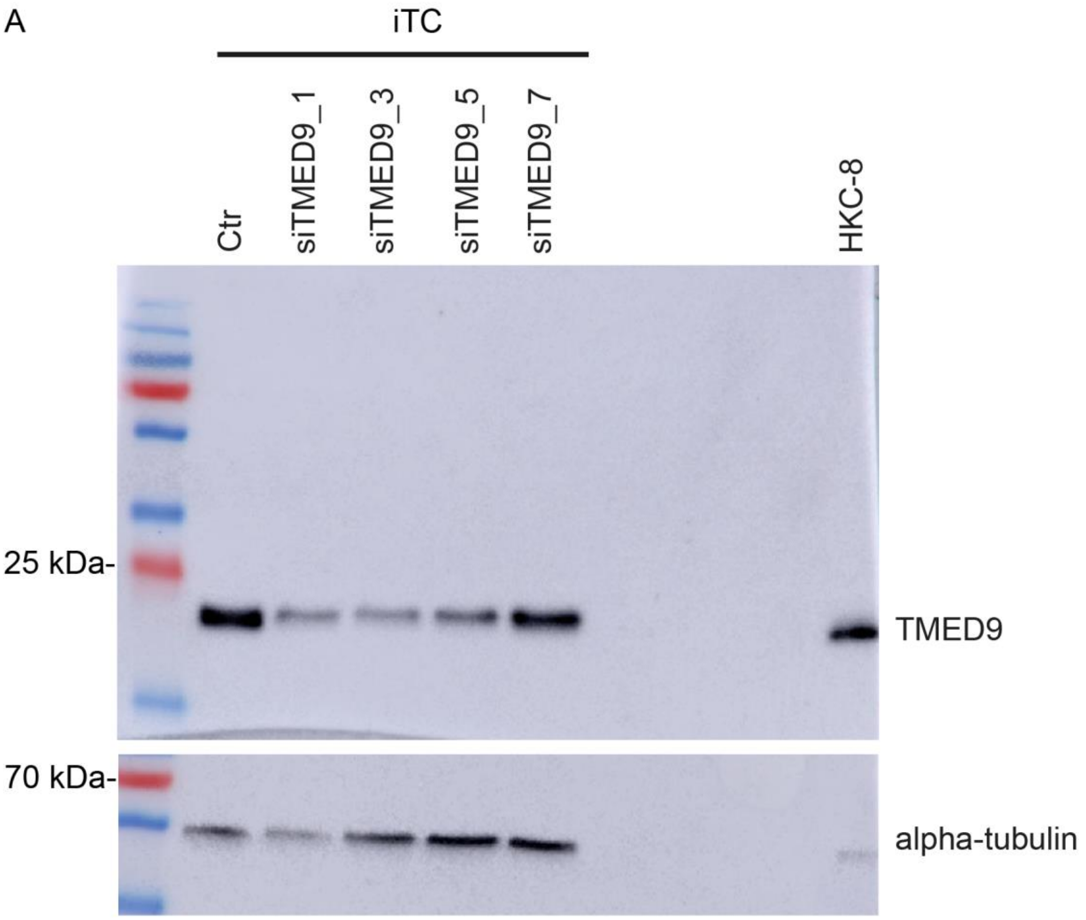

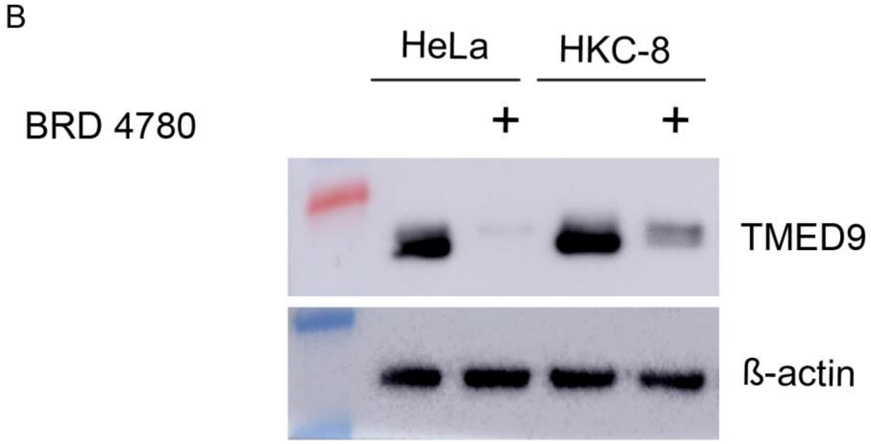

**Supplementary Figure S7:** *Evaluation of TMED9 knockdown and expression*

(A) Immunoblot of cell lysates from iTCs clone 03 (K03) of the affected individual ADTKD-0144 (A-30) for TMED9. iTCs were subjected to siRNA knockdown against TMED9 using 4 individual commercially available siRNAs (siTMED9_1, siTMED9-3, siTMED9_5 and siTMED9_7). Knockdown showed best results with siTMED9_1 and siTMED9_3, so these siRNAs were used in further experiments. Alpha-tubulin serves as loading control. (B) HeLa- and HKC-8 cells were subjected to BRD4780 treatment (10 µM, 24 h) and whole cell lysates were immunoblotted for TMED9. BRD4780 treatment strongly reduces TMED9 expression in both cell lines. ß-actin serves as loading control.

Supplementary Figure S8

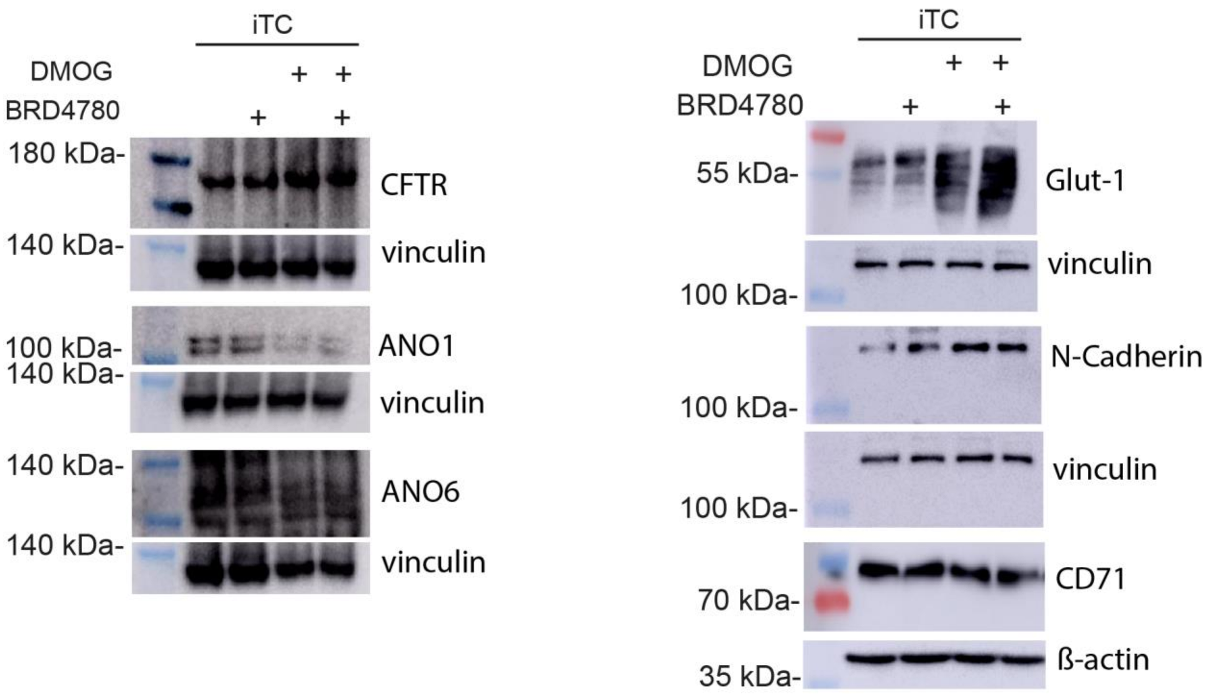

**Supplementary Figure S8:** *Proteins shuttling through the secretory pathway unaffected by BRD4780 treatment*

To analyze effects of BRD4780 on proteins apart from mucin 1 and MUC1-fs, iTCs from affected individual ADTKD-0150 (A-64) were treated with BRD4780 (10 μM) for 24 hours and DMOG (1 mM) to enhance transcriptional expression, where expected (e.g. HIF-target genes). Cell lysates where then subjected to immunoblotting against CFTR, ANO1, ANO6, Glut-1, N-Cadherin and CD71. These proteins were chosen, as they all shuttle through the secretory pathway during maturation in a similar matter as mucin 1 (and in part MUC1-fs). Protein expression of CFTR, Glut-1 and N-Cadherin is increased with DMOG treatment, yet all proteins analyzed do not show a reduction of expression after treatment with BRD4780. Vinculin and ß-actin serve as loading controls.

Supplementary Figure 9

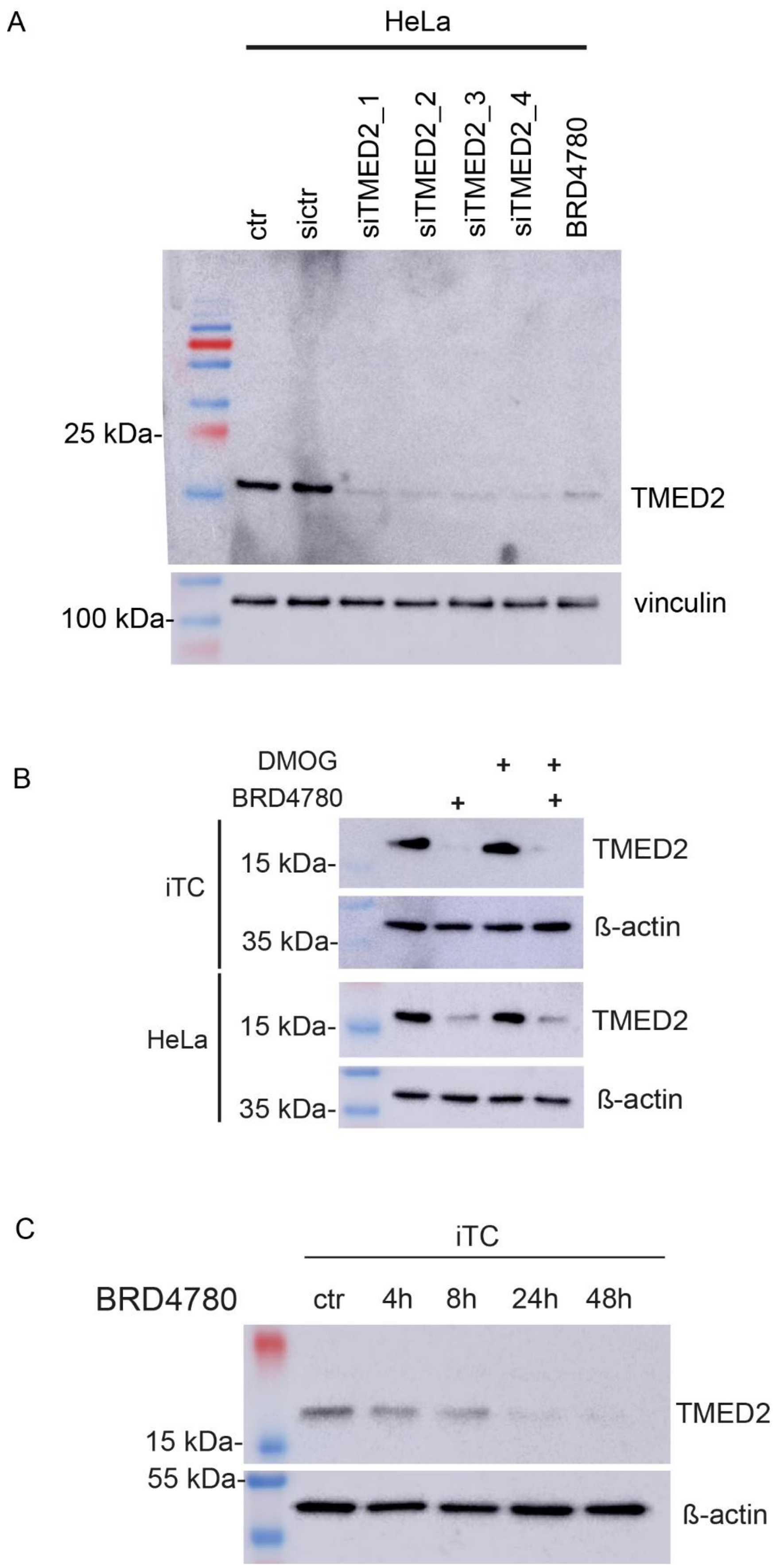

**Supplementary Figure S9:** *Evaluation of TMED2 antibody and response to BRD4780*

(A) To validate TMED2 antibody HeLa cells were subjected to siRNA knockdown using 4 individual, commercially available siRNAs (siTMED2_1, siTMED2_2, siTMED2_3 and siTMED2_4). Additionally, cells were treated with BRD4780 (10 µM) for 24 h. Cell lysates where the subjected to immunoblotting against TMED2. All four siRNAs, as well as BRD4780 treatment led to a clear downregulation of TMED2 protein. Vinculin serves as loading control. (B) Immunoblot for TMED2 was performed on cell lysates from iTCs from affected individual ADTKD-0150 (A-64) and from HeLa cells. Cells were treated with BRD4780 (10 µM) for 24 h. DMOG (1 mM) was added for 18 h. Both, iTCs and HeLa cells show a strong reduction of TMED2 protein levels after BRD4780 treatment. DMOG has no effect on TMED2. ß-actin serves as a loading control. (C) Immunoblot of cell lysates from iTC clone 03 (K03) of the affected individual ADTKD-0144 (A-30) for TMED2. Cells were treated with BRD4780 (10 µM) for 4h, 8h, 24h and 48 h compared to untreated control (ctr). A continuous decrease of TMED2 expression levels can be seen over time. Vinculin and ß-actin serve as loading controls.

Supplementary Figure S10

A

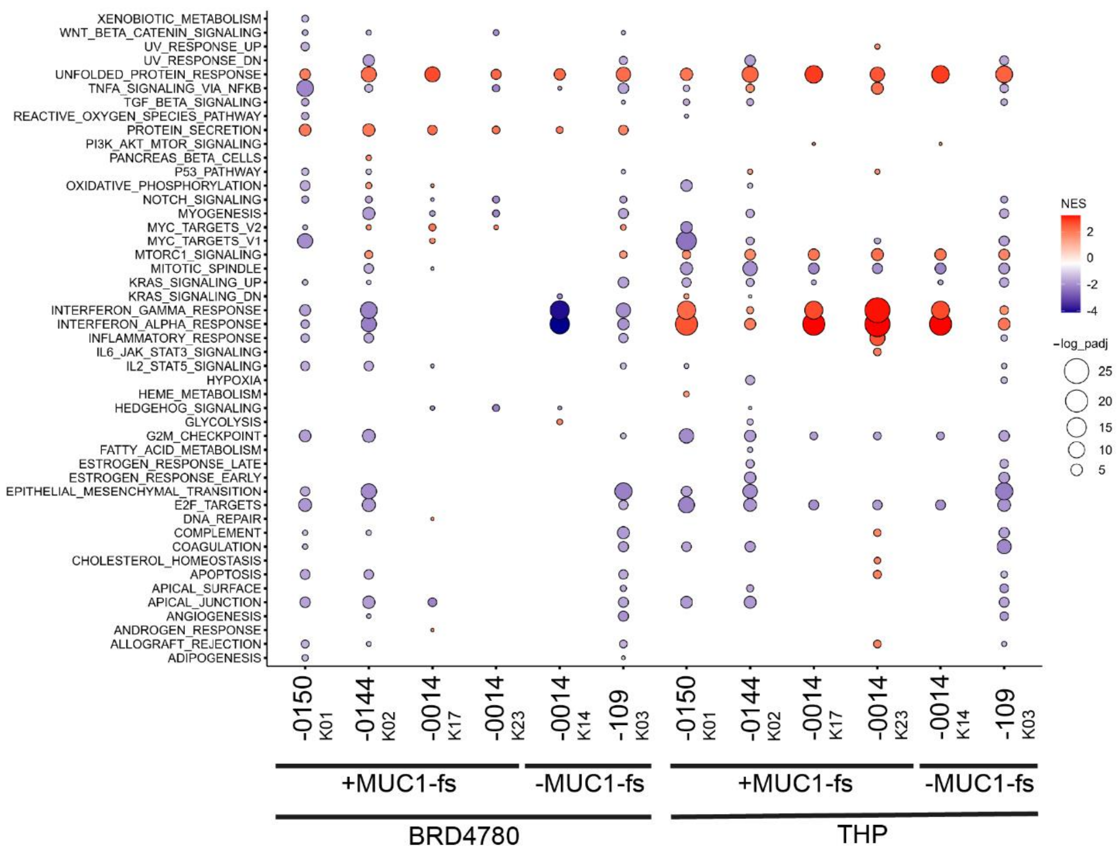

B

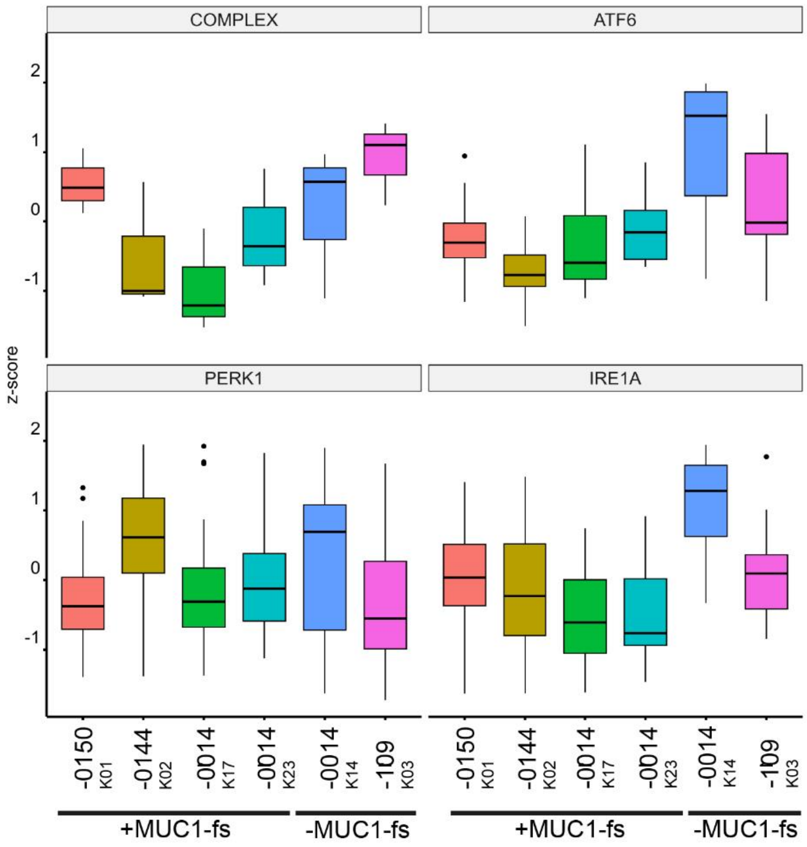

**Supplementary Figure S10:** *RNA-seq cellular pathway analyses*

(A) The bubble plot shows results of a Gene Set Enrichment Analysis (GSEA) using RNA-seq data from the indicated clones of cells and hallmark gene sets provided by MsigDB. Comprehensive RNA sequencing was conducted on immortalized tubular cells (iTCs) of five distinct clones (denoted as Kxx) originating from three individuals diagnosed with ADTKD-*MUC1*, specifically ADTKD-0150 (K01), ADTKD-0144 (K02), and ADTKD-0014 (K17 and K23). As negative controls, we incorporated a MUC1-fs negative clone (K14) sourced from ADTKD-0014 and an additional MUC1-fs negative clone (K03) from a healthy individual (UKER-109). These samples were subjected to RNA-seq analysis utilizing duplicate replicates. Cells were cultured under control conditions or treated with 10 µM BRD4780 for 24 h or 300 nM THP for 24 h and compared to controls. Differential expression analysis was conducted for each cell clone comparing either BRD4780 or THP treatment to control conditions. Enrichment of hallmark gene sets was determined by GSEA based on an adjusted p-value threshold of <0.05, employing the Benjamini-Hochberg (BH) correction method. Gene sets were subsequently visualized according to normalized enrichment scores (NES) and -log10(adjusted p-values). Red color indicates pathways that were induced by the respective treatment, while blue color depicting downregulated pathways. (B) RNA-seq data of the different clones of cells were analyzed for activation of the unfolded protein response using predefined marker genes(Adamson *et al*, 2016). Each box consists of genes assigned to either general UPR activation (complex) or specific UPR branches (ATF6, PERK1, IRE1A) as determined by(Adamson *et al.*, 2016). The z-score of all genes assigned to one branch was averaged and plotted as the mean value of the boxplot. RNA-seq was performed in technical replicates for each clone of cells. The mean transcripts per million (TPM) were calculated for the technical replicates of each cell clone associated with the assigned genes. Subsequently, the z-score for each gene within each cell clone was computed and visualized using a boxplot. RNA sequencing was conducted on the technical replicates for each cell clone.

**Supplementary Table S1: Antibodies (Ab) applied in our study**

| | | company (clone) | host | dilution IB | dilution IF |
|---|---|---|---|---|---|
| | **Primary Ab**: | | | | |
| 1 | MUC1-WT (1) (VNTR) | Cell Signaling (VU4H5), Danvers, USA | mouse | 1:1000 | 1:100 |
| 2 | MUC1-WT (2) (N-term) | Cell Signaling (D908K), Danvers, USA | rabbit | 1:1000 | |
| 3 | MUC1-fs (pAb3-fs) | n.a.(Knaup *et al*, 2018) | rabbit | 1:5000 | 1:1000 |
| 4 | TMED9 | Huabio (HA601086), Woburn, USA | mouse | 1:1000 | cells 1:500, tissue 1:50 |
| 5 | TMED9 | Proteintech (21620-1-AP), Rosemont, USA | rabbit | 1:1000 | Cells and tissue 1:50 |
| 6 | TMED2 | Proteintech (119-81-1-AP), Rosemont, USA | rabbit | 1:1000 | |
| 7 | CFTR | Alomone Labs (ACL-006) | rabbit | 1:500 | |
| 8 | ANO1 | Thermo (PA-87947) | rabbit | 1:1000 | |
| 9 | ANO6 | Generated by Davids Biotechnology | rabbit | 1:1000 | |
| 10 | Glut-1 | Alpha Diagnostics Intl Inc. (GT12-A), San Antonio, USA | rabbit | 1:1000 | |
| 11 | N-Cadherin | Santa Cruz Biotechnology, Inc. (sc-7939); Dallas, USA | rabbit | 1:1000 | |
| 12 | CD71 | Santa Cruz Biotechnology, Inc. (3B8 2A1), Dallas, USA | mouse | 1:500 | |
| 13 | β-Actin | Sigma (clone AC-15, 5441), Darmstadt, Germany | mouse | 1:25000 | |
| 14 | Vinculin | Novusbio (NB600-1293), Centennial, USA | mouse | 1:8000 | |
| 15 | α-Tubulin | Abcam (DM1A) | Mouse | 1:5000 | |
| | **Secondary Ab HRP:** | | **target/ host** | | |
| 16 | swine Anti-rabbit HRP | Dako (P0447), Santa Clara, USA | rabbit/ swine | 1:2000 | |
| 17 | goat Anti-mouse HRP | Dako (P0448), Santa Clara, USA | mouse/ goat | 1:2000 | |
| | **Secondary Ab Fluor:** | | | | |
| 18 | Alexa Fluor 594, Molecular Probes | Thermo Fisher Scientific (A28175), Darmstadt, Germany | mouse/ goat | | 1:500 |
| 19 | Alexa Fluor 594, Molecular Probes | Thermo Fisher Scientific (A11012), Darmstadt, Germany | rabbit/ goat | | 1:500 |
| 20 | Alexa Fluor 488, Molecular Probes | Thermo Fisher Scientific (A11070), Darmstadt, Germany | rabbit/ goat | | 1:500 |
| 21 | Alexa Fluor 488, Molecular Probes | Thermo Fisher Scientific (A28175), Darmstadt, Germany | mouse/ goat | | 1:500 |

Dilutions for immunoblotting (IB) and Immunofluorescence (IF), as indicated.

**Supplementary Table S2: siRNAs applied in our study**

| | Name | Company | Catalog no. | Target sequence 5‘->3‘ |
|---|---|---|---|---|
| 1 | Hs_MUC1_5 | Qiagen | SI00162988 | CTGCAGAGAGACATTTCTGAA |
| 2 | Hs_MUC1_10 | Qiagen | SI04949826 | CAGCACCGACTACTACCAAGA |
| 3 | Hs_TMED9_1 | Qiagen | SI00746879 | CACCTCAGAATCACAGTGTTA |
| 4 | Hs_TMED9_3 | Qiagen | SI00746893 | CCGGACGCAGCTGTATGACAA |
| 5 | Hs_TMED9_5 | Qiagen | SI04303894 | CCAGCCATACCTGTTCTGGAA |
| 6 | Hs_TMED9_6 | Qiagen | SI04338880 | CAGGTAGGTGAACATGCCAAT |
| 7 | Hs_TMED2_1 | Qiagen | SI04151980 | TAGGTCCTTCCAGGAACTCAA |
| 8 | Hs_TMED2_2 | Qiagen | SI04171804 | CTCGGGCTATTTCGTTAGCAT |
| 9 | Hs_TMED2_3 | Qiagen | SI04172070 | CAGTATGAATCTTGACGGTTT |
| 10 | Hs_TMED2_4 | Qiagen | SI04184894 | CAGGTTCGGTAAATAAACAAT |
| 11 | Negative Control siRNA | Qiagen | 1027310 | AATTCTCCGAACGTGTCACGT |
